# Proteomic Discovery of Urinary Myoglobin as a Noninvasive Biomarker for PROCHOB caused by *CUBN* Variants

**DOI:** 10.64898/2026.03.26.26349155

**Authors:** Yuta Inoki, Tomoko Horinouchi, Nana Sakakibara, Shinya Ishiko, Asahi Yamamoto, Shuhei Aoyama, Yuka Kimura, Yuta Ichikawa, Yu Tanaka, Atsushi Kondo, Tomohiko Yamamura, Shingo Ishimori, Yoshinori Araki, Takako Asano, Junya Fujimura, Shuichiro Fujinaga, Riku Hamada, Natsumi Inoue, Hiroshi Kaito, Kyoko Kiyota, Anna Kobayashi, Yasuko Kobayashi, Naonori Kumagai, Hiroki Miyano, Yoshiyuki Ohtomo, Satoshi Sasaki, Ryota Suzuki, Mami Washio, Yuko Yamada, Yoko Yamasaki, Tadafumi Yokoyama, Kazumoto Iijima, China Nagano, Kandai Nozu

## Abstract

Chronic benign proteinuria (PROCHOB)—caused by biallelic pathogenic variants in *CUBN*—presents in childhood as isolated, asymptomatic tubular proteinuria with preserved long-term kidney function. Because its clinical presentation closely mimics early stage glomerular diseases with moderate proteinuria and without increased urinary β2-microglobulin (uBMG) and α1-microglobulin, numerous patients undergo unnecessary kidney biopsies and receive angiotensin-converting enzyme inhibitors or angiotensin II receptor blockers before genetic testing is considered. Using high-throughput aptamer-based urinary proteomics (SomaScan®), we identified urinary myoglobin as a disease-specific biomarker for PROCHOB. We developed and confirmed a diagnostic approach in which the urinary myoglobin-to-creatinine (uMB/Cr) ratio robustly distinguishes PROCHOB from other moderate glomerular proteinuric kidney diseases. Although certain cases of Dent disease causing megalin dysfunction exhibit increased urinary myoglobin levels, PROCHOB and Dent disease can be clearly distinguished based on the uBMG-to creatinine ratio. This biomarker reflects impaired proximal tubular protein reabsorption because of cubilin dysfunction and remains normal in healthy individuals or those with typical glomerular diseases with moderate proteinuria. Our findings establish a noninvasive diagnostic tool for PROCHOB that prompts targeted genetic testing for *CUBN* variants using the uMB/Cr and urinary uBMG-to-creatinine ratios. This strategy has the potential to transform the clinical diagnostic pathway for isolated proteinuria.

## Introduction

Most proteins that traverse the glomerular capillary wall are reabsorbed in the proximal tubule through receptor-mediated endocytosis, resulting in minimal urinary protein excretion in healthy individuals. This process is primarily mediated by the endocytic receptors megalin and cubilin that function cooperatively (1). Specific filtered proteins are reabsorbed predominantly through either megalin- or cubilin-dependent pathways, whereas certain ligands use both receptors (1, 2). Cubilin, encoded by the *CUBN* gene, is a 460 kDa glycoprotein that binds albumin, hemoglobin, myoglobin, transferrin, the intrinsic factor–vitamin B12 complex, apolipoprotein A-I, and other low-molecular-weight proteins (1–8).

Biallelic pathogenic variants in *CUBN* are classically known to cause Imerslund–Gräsbeck syndrome (OMIM #261100), a rare disorder characterized by selective vitamin B12 malabsorption and low-molecular-weight tubular proteinuria (9). Recently, these variants located in the C-terminal region of *CUBN* have been reported to cause persistent isolated tubular proteinuria without hematologic abnormalities or vitamin B12 deficiency (10–12). This condition is now recognized as a distinct clinical entity, termed chronic benign proteinuria (PROCHOB; OMIM #618884). Pathophysiologically, PROCHOB is a tubular disorder resulting from impaired cubilin-mediated reabsorption, including albumin in the proximal tubule. However, unlike typical tubular proteinuria, PROCHOB often lacks significant increases in conventional low-molecular-weight protein markers, such as urinary β2-microglobulin (uBMG) or α1-microglobulin (11–14). Consequently, PROCHOB clinically presents with isolated albuminuria that closely resembles progressive glomerular diseases, despite being associated with preserved long-term kidney function, even without treatment (11, 13). Owing to this glomerular-like (rather than truly glomerular) profile, standard therapies for glomerular proteinuria, such as angiotensin-converting enzyme inhibitors (ACE-I) or angiotensin II receptor blockers (ARB), are generally ineffective and unnecessary (11–13). In Japanese cohorts, our group has shown that many numerous PROCHOB cases have been detected through routine urine screening at 3 years of age, and kidney prognosis is generally favorable (12). Because kidney biopsy findings are typically yield normal or non-specific, a definitive diagnosis relies solely on genetic testing.

This unique biochemical profile makes it exceptionally challenging to clinically differentiate between PROCHOB and early-stage progressive glomerular diseases. Although previous studies have emphasized the significance of genetic testing for a definitive diagnosis (11, 13, 15, 16), its limited accessibility in routine practice often results in unnecessary invasive kidney biopsies or inappropriate ACE-I/ARB therapy (11–13). Because of the potential side effects of pharmacologic therapy and invasiveness of biopsy, there is an urgent clinical need for a non-invasive biomarker specific to PROCHOB. A similar diagnostic paradigm exists for Dent disease. Pathogenic variants in *CLCN5* (type1) or *OCRL* (type 2) disrupt proximal tubular megalin- and cubilin-mediated receptor endocytosis, causing impaired reabsorption of filtered low-molecular-weight proteins (17, 18). In this context, uBMG is typically significantly increased and serves as a clinically useful biomarker. This raises suspicion for Dent disease and helps prioritize genetic testing over kidney biopsy, thereby reducing unnecessary biopsies (19). Recently, SomaScan^®^ proteomics—an aptamer-based platform for high-sensitivity and high-throughput protein quantification—has been increasingly used for biomarker discovery in various kidney diseases (20–23). However, to date, no biomarkers have yet been identified that can specifically screen for PROCHOB.

This study aimed to identify practical urinary biomarkers to support the diagnosis of PROCHOB. We performed urinary proteomic screening using SomaScan and assessed the diagnostic utility of the selected marker in an independent validation cohort. We aimed to establish a clinically applicable biomarker that can prompt earlier genetic testing, thereby enhancing diagnostic efficiency and reducing unnecessary interventions, such as ACE-I/ARB therapy and kidney biopsy.

## Results

### Somascan analysis

We compared the urinary protein profiles of seven patients with genetically confirmed PROCHOB, seven with INS, and six with genetically confirmed Alport syndrome. Detailed patient data are provided in Supplementary Table 1. Compared with the combined comparator group (INS and Alport), 23 proteins in the PROCHOB group met the pre-specified nomination criteria (Table 1). Among them, only myoglobin and α-amylase 1 were known ligands of cubilin/megalin. Following FDR correction across all 7,288 proteins, only dipeptidyl peptidase 4 (DPP4) remained significant (q < 0.05). Because DPP4 is not a known ligand of cubilin/megalin and may reflect processes unrelated to receptor-mediated proximal tubular reabsorption, we did not pursue further validation. Urinary amylase activity exceeded the normal range in only one of the seven patients with PROCHOB (Supplementary Table 2); therefore, subsequent validation analyses focused on urinary myoglobin.

**Table 1.**
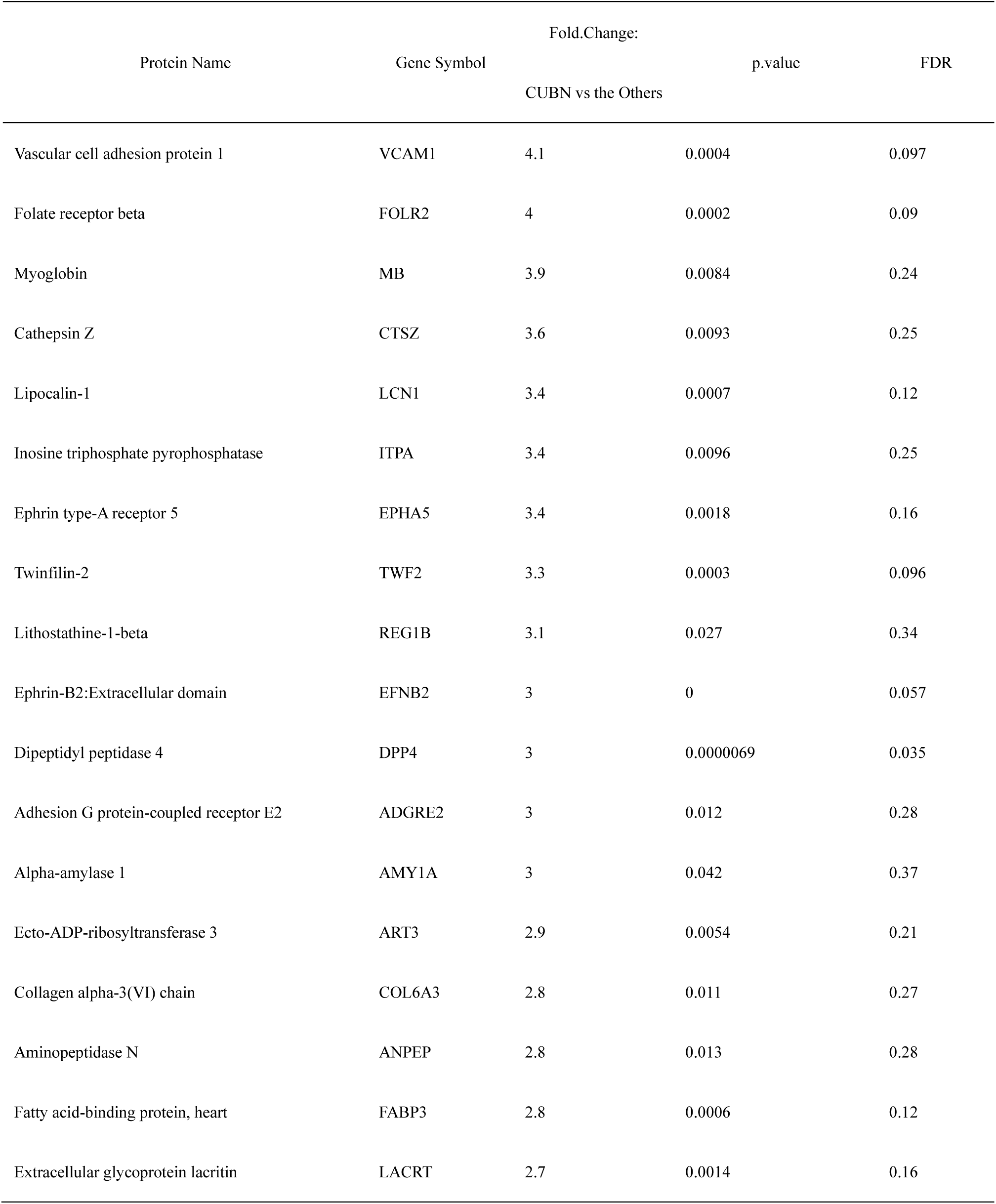
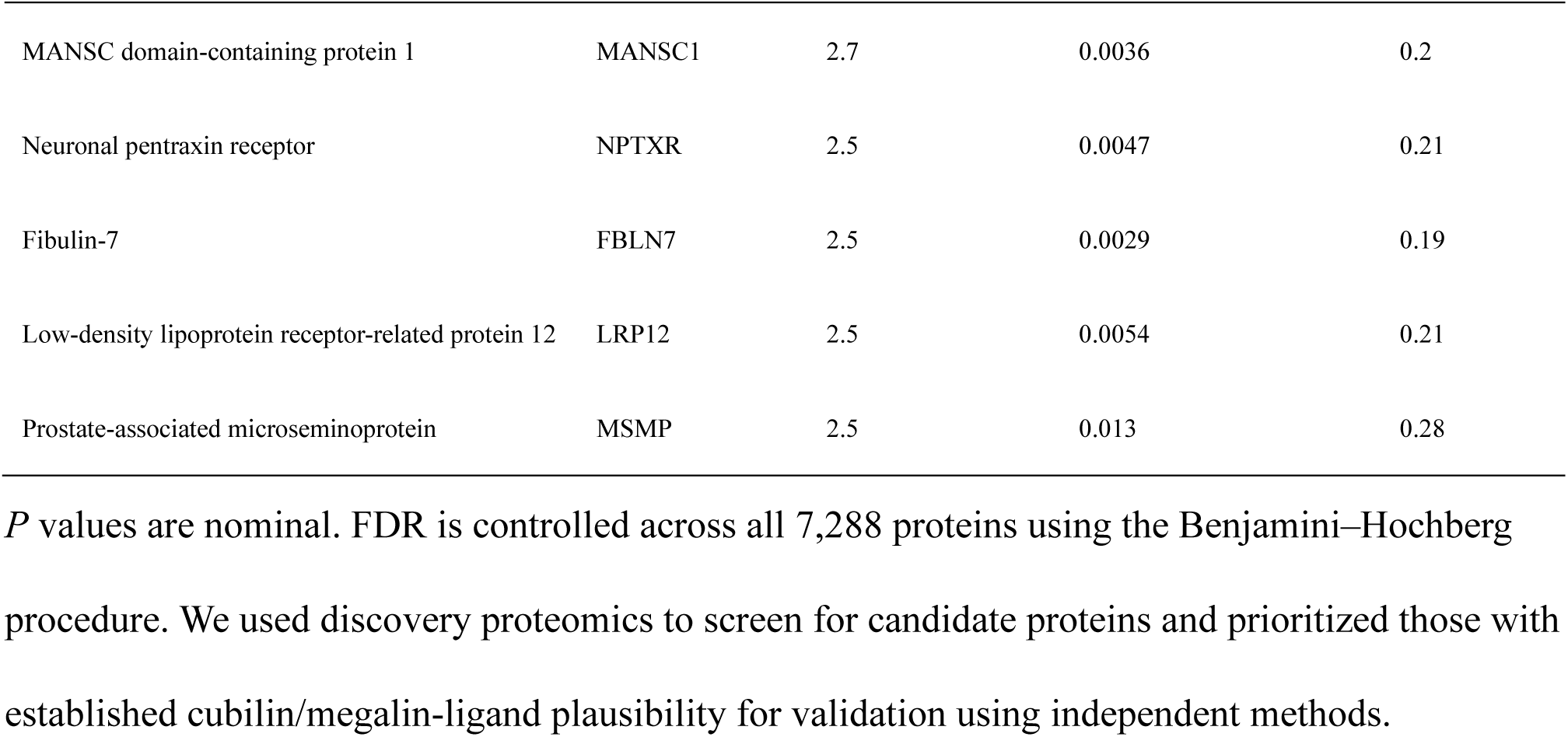
Upregulated proteins in chronic benign proteinuria (PROCHOB) compared with Alport syndrome and idiopathic nephrotic syndrome in the SomaScan discovery analysis.

We assessed other cubilin-mediated tubular reabsorption proteins. However, none of these proteins exhibited a significant increase in urinary levels in the PROCHOB group (Supplementary Table 3).

### Myoglobin Validation Cohort

#### Patient Characteristics

Urinary myoglobin was measured in 25 patients from 21 families with genetically confirmed PROCHOB, including eight patients who also participated in our previous cohort study (12). Variant-level details and in silico predictions are provided in Supplementary Table 4. For comparison, 33 patients were included in the moderate proteinuric glomerular disease group (0.15 < urinary total protein-to-creatinine [uTP/Cr] ≤ 4.0 g/gCr), comprising 11 with asymptomatic proteinuria, eight with IgA vasculitis nephritis, seven with IgA nephropathy, three with Alport syndrome, two with hereditary nephrotic syndrome, one with anti-neutrophil cytoplasmic antibody-associated vasculitis, and one with C3 nephritis. Among the 11 patients with asymptomatic proteinuria, four had orthostatic proteinuria, four had partial remission during prednisolone treatment for idiopathic nephrotic syndrome (INS) (0.15 < uTP/Cr < 2.0 g/gCr), and three had persistent proteinuria without pathogenic variants on genetic testing. Additionally, 11 patients were included in the severe proteinuric glomerular disease group (uTP/Cr > 4.0 g/gCr), comprising seven with INS, three with IgA vasculitis nephritis, and one with hereditary nephrotic syndrome. Moreover, six patients with Dent disease and 10 proteinuria-negative controls were enrolled. Detailed clinical data for each patient are provided in Supplementary Table 5.

The baseline clinical characteristics are summarized in Table 2. In the PROCHOB group, most patients were identified through routine urinalysis screening (24/25, 96.0 %), and none presented with symptoms, such as edema. The median age at detection was 3.0 years (IQR 3.0–3.5), and the median age at laboratory testing was 10.0 years (IQR 6.1–14.6). Creatinine-based estimated glomerular filtration rate (eGFR) was preserved across groups, whereas serum albumin levels were lower in the severe proteinuric glomerular disease group, consistent with the presence of nephrotic-range proteinuria. Occult hematuria occurred in 24.0 % (6/25) of patients in the PROCHOB groupand in 72.7 % (24/33) of patients in the moderate proteinuric glomerular disease group. Notably, all patients with PROCHOB exhibiting positive occult hematuria had <1 RBC/high-power field on urine sediment microscopy.

**Table 2.** Baseline characteristics of the five study groups.

| Variable | Chronic benign<br>proteinuria<br>(PROCHOB)<br>(n = 25) | Moderate proteinuric<br>glomerular diseases<br>(n = 33) | Severe<br>proteinuric<br>glomerular<br>diseases<br>(n = 11) | Dent disease<br>(n = 6) | Proteinuria-negative<br>controls<br>(n = 10) |
| --- | --- | --- | --- | --- | --- |
| Age at detection (years) | 3.0 (3.0–3.5) | 6.0 (3.0–10.5) | 10.1 (2.9–13.6) | 4.5 (2.5–8.3) | 3.8 (3.0–12.5) |
| Age at laboratory testing (years) | 10.0 (6.1–14.6) | 11.9 (5.7–13.7) | 10.4 (5.7–14.1) | 10.9 (8.5–13.1) | 11.5 (4.0–13.9) |
| Male sex, n (%) | 14 (56.0) | 17 (51.5) | 8 (72.7) | 5 (83.3) | 6 (60.0) |
| Clinical presentation at initial<br>detection, n (%) |  |  |  |  |  |
| – Routine urinalysis screening | 24 (96.0) | 20 (60.6) | 0 (0.0) | 5 (83.3) | 6 (60.0) |
| – Incidental urinalysis | 1 (4.0) | 0 (0.0) | 0 (0.0) | 1 (16.7) | 0 (0.0) |
| – Macroscopic hematuria | 0 (0.0) | 1 (3.0) | 0 (0.0) | 0 (0.0) | 0 (0.0) |
| – Edema | 0 (0.0) | 4 (12.1) | 8 (72.7) | 0 (0.0) | 0 (0.0) |
| – Purpura | 0 (0.0) | 8 (24.2) | 3 (27.3) | 0 (0.0) | 0 (0.0) |
| – Others | 0 (0.0) | 0 (0.0) | 0 (0.0) | 0 (0.0) | 4 (40.0) |
| Laboratory findings |  |  |  |  |  |
| Cr-eGFR (mL/min/1.73 m<sup>2</sup>) | 114.0 (99.2–137.3) | 123.3 (105.6–140.0) | 123.0 (110.8–<br>144.2) | 124.3 (68.6–<br>145.5) | 114.8 (101.6–<br>128.4) |
| sAlb (g/dL) | 4.5 (4.1–4.6) | 4.0 (3.4–4.3) | 2.7 (2.3–3.3) | 4.6 (4.4–4.9) | 4.5 (4.3–4.7) |
| Urinary occult blood, n (%) | 6 (24.0) | 24 (72.7) | 4 (36.4) | 0 (0.0) | 4 (40.0) |
| Kidney biopsy performed, n (%) | 17 (68.0) | 20 (60.6) | 6 (54.6) | 0 (0.0) | 0 (0.0) |
| Pathological findings, n (%) |  |  |  |  |  |
| – Minimal glomerular abnormalities | 17 (100) | 14 (70.0) | 5 (83.3) | 0 (0.0) | 0 (0.0) |
| – Focal segmental<br>glomerulosclerosis | 0 (0.0) | 1 (5.0) | 1 (16.7) | 0 (0.0) | 0 (0.0) |
| – Diffuse mesangial proliferation | 0 (0.0) | 5 (25.0) | 0 (0.0) | 0 (0.0) | 0 (0.0) |

Data are presented as median (IQR) or n (%), as appropriate. The moderate proteinuric glomerular disease group is defined as $0.15 < \text{uTP/Cr} \leq 4.0 \text{ g/gCr}$ , and the severe proteinuric glomerular disease group as $\text{uTP/Cr} > 4.0 \text{ g/gCr}$ . First symptom presentation indicates the clinical context that results in the initial recognition of abnormal urinalysis findings. Cr-eGFR, creatinine-based estimated glomerular filtration rate; sAlb, serum albumin; and uTP/Cr, urinary total protein-to-creatinine ratio.

Kidney biopsy was performed in 68.0 (17/25) and 60.6 % (20/33) of patients in the PROCHOB and moderate proteinuric glomerular disease groups, respectively. Among all patients in the PROCHOB group, minimal glomerular abnormalities (17/17, 100 %) were observed. In the moderate proteinuric glomerular disease group, the pathological diagnoses included minimal glomerular abnormalities (14/20, 70.0 %), focal segmental glomerulosclerosis (1/20, 5.0 %), and diffuse mesangial proliferation (5/20, 25.0 %).

Before a definitive genetic diagnosis, 11 patients (44.0 %) in the PROCHOB group received ACE-Iand/or ARBs for proteinuria. However, no clinically meaningful reduction in proteinuria was observed, and treatment was discontinued after genetic confirmation.

#### Urinary Findings Including Urinary Myoglobin Levels Across Patient Groups

Table 3 summarizes uTP/Cr, uBMG, urinary myoglobin, and urinary myoglobin-to-creatinine ratio (uMB/Cr) across the five predefined groups. The uTP/Cr ratio differed across groups (Kruskal–Wallis, p < 0.0001; Fig. 1A), with the highest values observed in the severe proteinuric glomerular disease group. The uTP/Cr ratio was comparable between the PROCHOB and moderate proteinuric glomerular disease groups (Mann–Whitney U, p = 0.47; Fig. 2A). uBMG levels differed across groups and were significantly increased in Dent disease (Kruskal–Wallis, p < 0.001; Fig. 3A). In contrast, uBMG levels were comparable between the PROCHOB and moderate proteinuric glomerular disease groups (Mann–Whitney U, p = 0.20; Fig. 3B). Similarly, the uBMG/Cr ratio differed across the five groups (Kruskal–Wallis, p < 0.001; Fig. 3C) and was modestly higher in the PROCHOB group than that in the moderate proteinuric glomerular disease group (Mann–Whitney U, p = 0.028; Fig. 3D).

**Figure 1.**
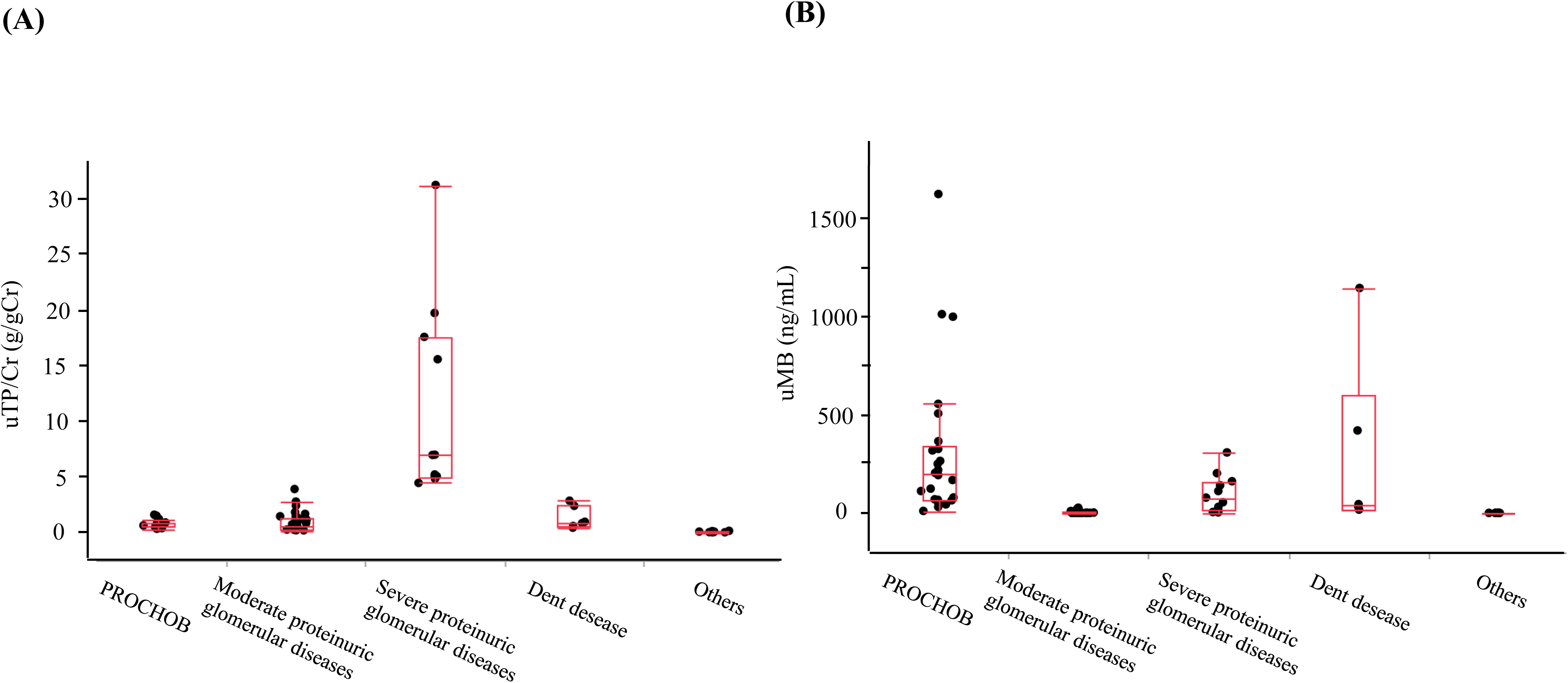
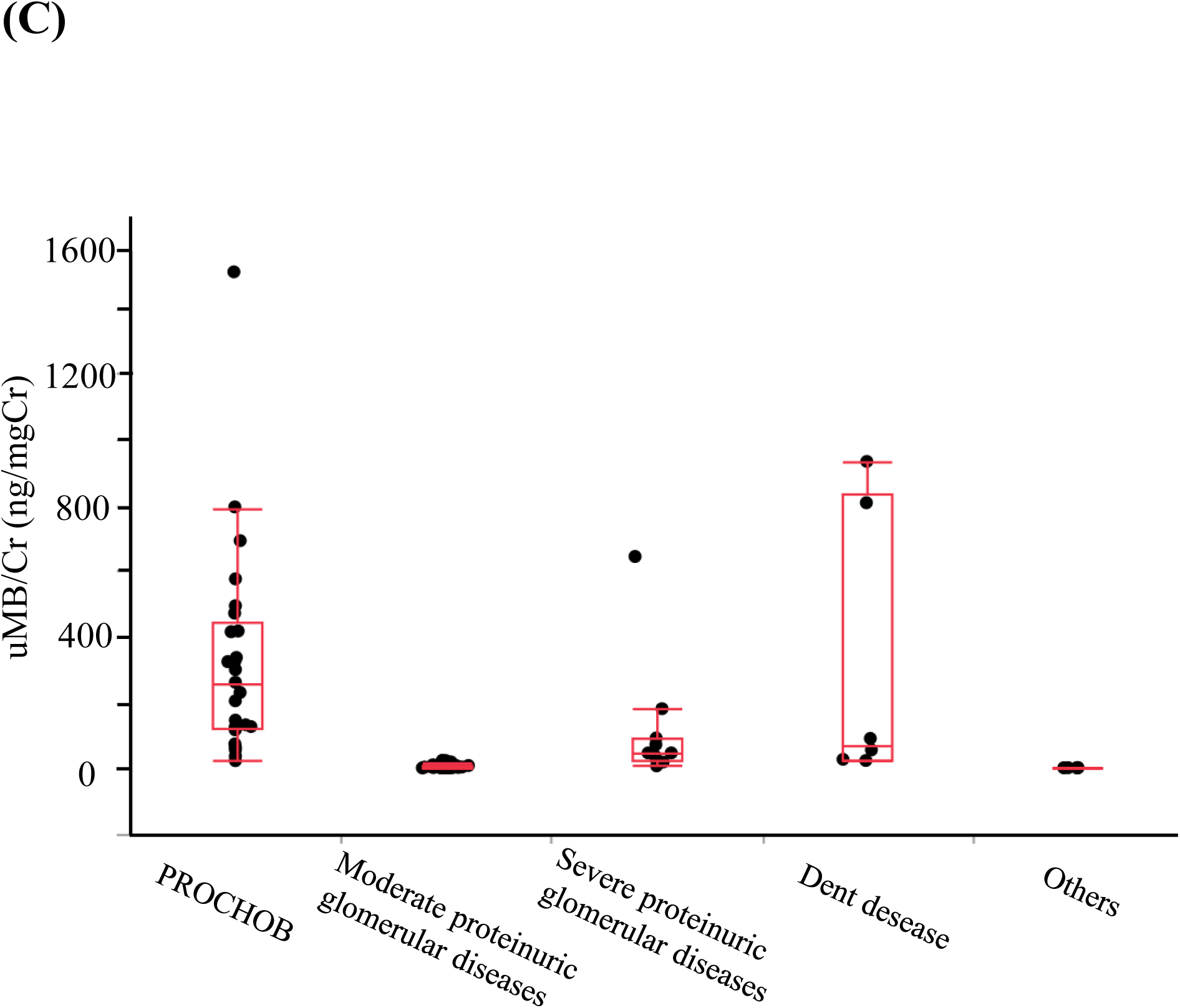
Urinary myoglobin (uMB), myoglobin-to-creatinine (uMB/Cr) ratio, and proteinuria across the five predefined study groups. (A) uMB (ng/mL). (B) uMB/Cr (ng/mgCr). (C) Urine protein-to-creatinine ratio (uTP/Cr, g/gCr). Groups include chronic benign proteinuria (PROCHOB) (n = 24), moderate proteinuric glomerular diseases (0.15 < uTP/Cr ≤ 4.0 g/gCr; n = 33), severe proteinuric glomerular diseases (uTP/Cr > 4.0 g/gCr; n = 11), Dent disease (n = 6), and proteinuria-negative controls (Others; n = 10). Box-and-whisker plots demonstrate the median (center line) and IQR (box). Whiskers indicate 1.5× the IQR. Each dot represents an individual participant. Overall differences among the groups are assessed using the Kruskal–Wallis test (uMB, p < 0.0001; uMB/Cr, p < 0.0001; and uTP/Cr, p < 0.0001). Pairwise post hoc comparisons are performed using Dunn’s test with Bonferroni correction. The uMB and uMB/Cr ratios are higher in the PROCHOB, severe proteinuric glomerular disease, and Dent disease groups than that in the moderate proteinuric glomerular disease group (all adjusted p < 0.05). Proteinuria-negative controls had lower uMB and uMB/Cr than that of the PROCHOB and Dent disease groups (adjusted p < 0.01 for uMB and adjusted p < 0.01 for uMB/Cr).

**Figure 2.**
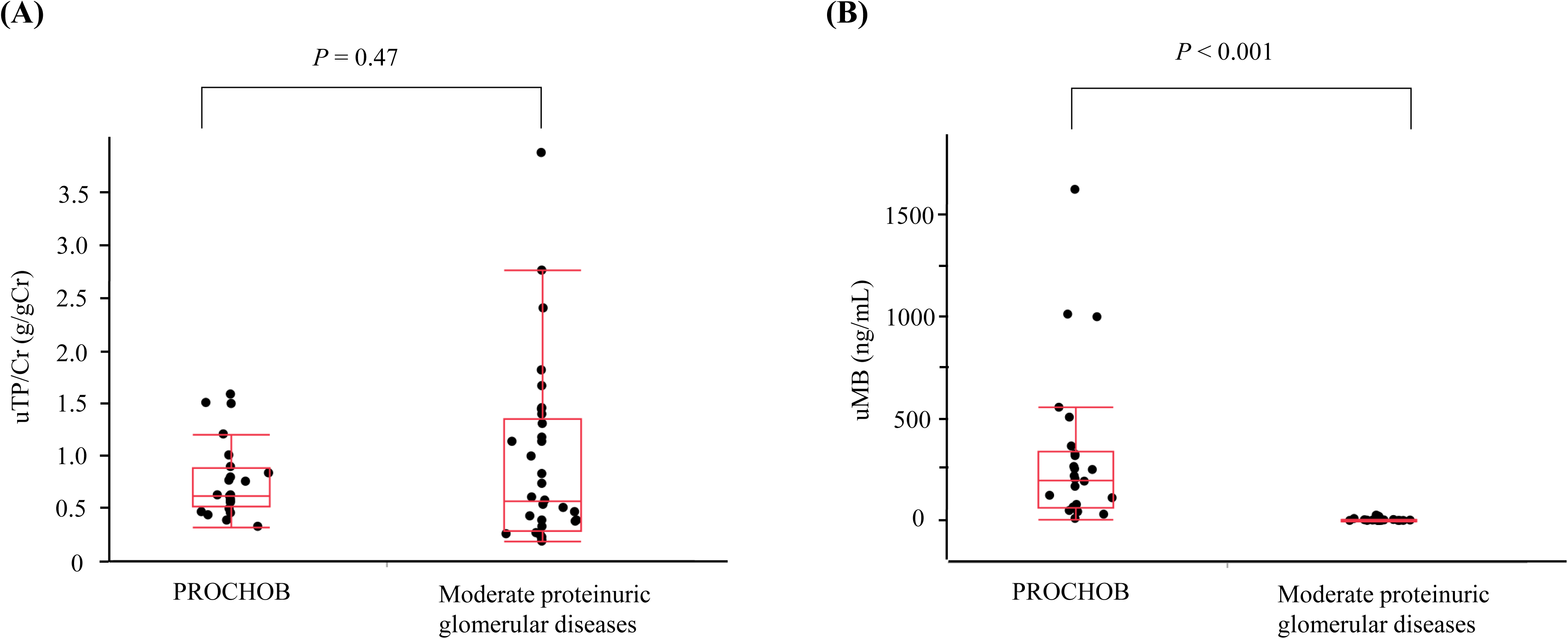
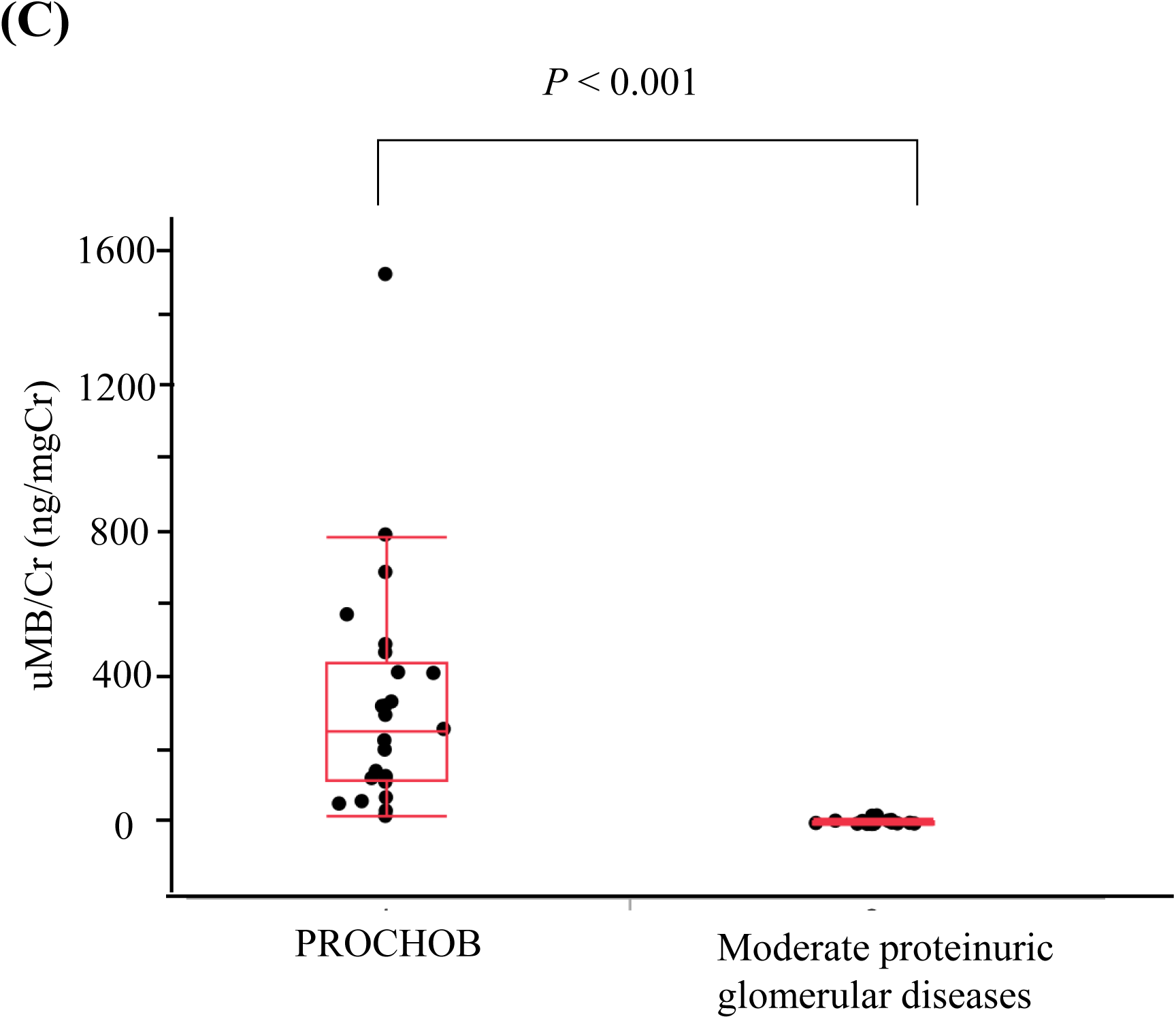
Comparison of urinary myoglobin (uMB), myoglobin-to-creatinine ratio (uMB/Cr), and proteinuria between chronic benign proteinuria (PROCHOB) and moderate proteinuric glomerular diseases. (A) uMB (ng/mL). (B) uMB/Cr (ng/mgCr). (C) Urinary total protein-to-creatinine (uTP/Cr) (g/gCr). PROCHOB (n = 24) is compared with moderate proteinuric glomerular diseases (0.15 < uTP/Cr ≤ 4.0 g/gCr; n = 33). Box-and-whisker plots demonstrate the median (center line) and IQR (box). Whiskers indicate 1.5× the IQR. Each dot represents an individual participant. Group differences are assessed using the Mann–Whitney U test (uMB, p < 0.001; uMB/Cr, p < 0.001; and uTP/Cr, p = 0.47).

**Figure 3.**
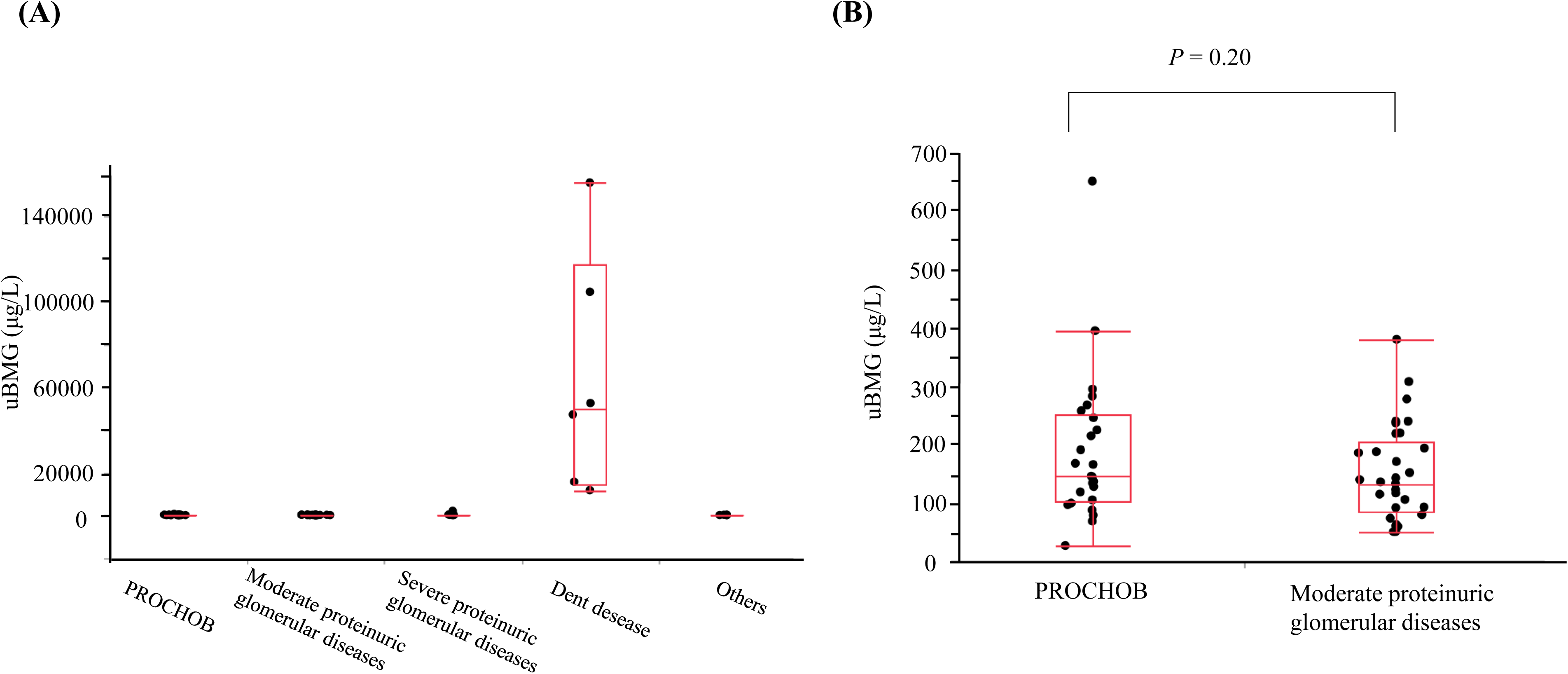
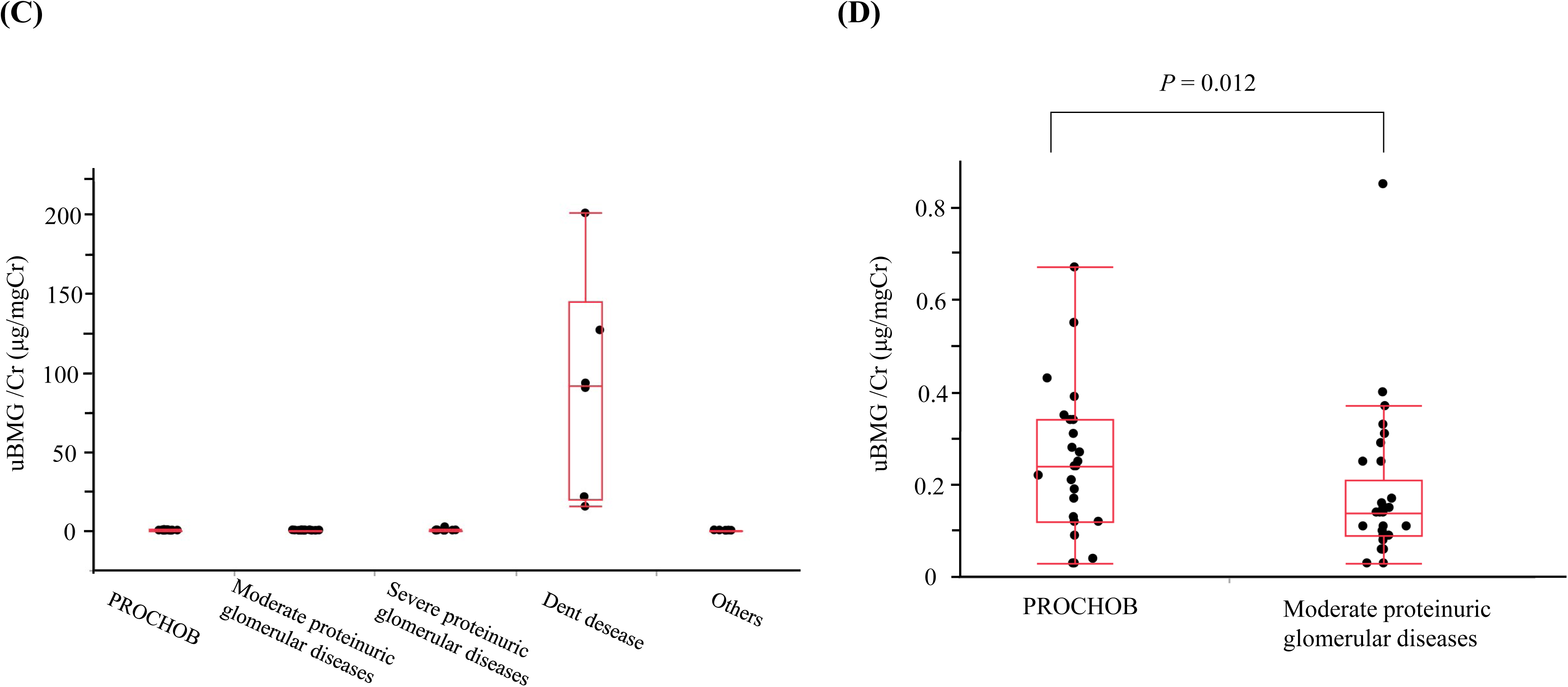
Urinary β2-microglobulin (uBMG) concentration and β2-microglobulin-to-creatinine (uBMG /Cr) ratio across the five predefined groups and comparison between chronic benign proteinuria (PROCHOB) and moderate proteinuric glomerular diseases. (A) uBMG (µg/L) across the five groups (PROCHOB, moderate proteinuric glomerular diseases, severe proteinuric glomerular diseases, Dent disease, and others) (Kruskal–Wallis, p < 0.001). (B) Direct comparison of uBMG between PROCHOB and moderate proteinuric glomerular diseases (Mann–Whitney U, p = 0.20). (C) uBMG /Cr (µg/mgCr) across the five groups (Kruskal–Wallis, p < 0.001). (D) Direct comparison of uBMG /Cr between PROCHOB and moderate proteinuric glomerular diseases (Mann–Whitney U, p = 0.028). Box-and-whisker plots demonstrate the median (center line) and IQR (box). Whiskers indicate 1.5× the IQR. Each dot represents an individual participant.

**Table 3.** Urinary myoglobin and related laboratory findings across five study groups.

| Variable | Chronic benign proteinuria<br>(PROCHOB) (n = 25) | Moderate proteinuric<br>glomerular diseases (n = 33) | Severe proteinuric<br>glomerular diseases<br>(n = 11) | Dent disease<br>(n = 6) | Proteinuria-negative controls<br>(n = 10) |
| --- | --- | --- | --- | --- | --- |
| uMB positive (> 2.0 ng/mL) | 25/25 (100 %) | 18/33 (54.6 %) | 11/11 (100 %) | 6/6 (100%) | 0/10 (0%) |
| uMB (ng/mL) | 204.0 (68.5–345.5) | 2.2 (1.0–5.2) | 78.7 (16.4–161.5) | 38.7 [18.9–600.4] | 1.0 [1.0–1.0] |
| uMB/Cr (ng/mgCr) | 260.5 (120.3–443.6) | 2.5 (1.1–7.9) | 45.8 (23.7–90.9) | 72.4 [25.4–837.8] | 1.1 [0.7–1.3] |
| uBMG (µg/L) | 145.0 (101.7–251.0) | 132.0 (85.0–205.5) | 236.0 (126.0–338.0) | 49742.5 [14700.3–<br>117107.3] | 112.5 [81.5–174.3] |
| uBMG /Cr (µg/mgCr) | 0.24 (0.12–0.34) | 0.14 (0.09–0.21) | 0.29 (0.12–0.43) | 91.7 [19.9–145.2] | 0.13 [0.05–0.23] |
| uTP/Cr (g/gCr) | 0.62 (0.52–0.89) | 0.57 (0.29–1.35) | 6.93 (5.00–17.50) | 0.89 [0.53–2.50] | 0.05 [0.02–0.08] |

Data are presented as median (IQR) unless otherwise specified. The moderate proteinuric glomerular disease group was defined as $0.15 < \text{uTP/Cr} \leq 4.0$ g/gCr, and the severe proteinuric glomerular disease group as $\text{uTP/Cr} > 4.0$ g/gCr. uMB positive is defined as urinary myoglobin > 2.0 ng/mL that corresponds to the assay lower limit of quantification (LLOQ). Values below the LLOQ are imputed as 1.0 ng/mL for analyses. uTP/Cr, urinary total protein-to-creatinine ratio; uBMG, urinary β2-microglobulin; uBMG /Cr, urinary β2-microglobulin-to-creatinine ratio; uMB, urinary myoglobin; and uMB/Cr, urinary myoglobin-to-creatinine ratio

Urinary myoglobin differed across the five groups (Kruskal–Wallis, p < 0.0001; Fig. 1B). The median myoglobin concentrations were 204.0 (IQR 68.5–345.5), 2.2 (IQR 1.0–5.2), 78.7 (IQR 16.4–161.5), 38.7 (IQR 18.9–600.4), and 1.0 ng/mL (IQR 1.0–1.0) in the PROCHOB, moderate proteinuric glomerular disease, severe proteinuric glomerular disease, Dent disease, and proteinuria-negative control groups, respectively. Post-hoc Dunn’s test with Bonferroni correction exhibited higher urinary myoglobin levels in the PROCHOB, severe proteinuric glomerular disease, and Dent disease groups than that in the moderate proteinuric glomerular disease group (all adjusted p < 0.05). Additionally, urinary myoglobin was higher in the PROCHOB and Dent disease groups than that in the proteinuria-negative group (adjusted p < 0.01). Consistent with the clinically most relevant comparison, myoglobin levels were higher in the PROCHOB group than that in the moderate proteinuric glomerular disease group (Mann–Whitney U test, p < 0.001; Fig. 2B).

The uMB/Cr levels differed across the five groups (Kruskal–Wallis, p < 0.0001; Fig. 1C). The median uMB/Cr ratios were 260.5 (IQR 120.3–443.6), 2.5 (IQR 1.1–7.9), 45.8 (IQR 23.7–90.9), 72.4 (IQR 25.4–837.8), and 1.1 ng/mgCr (IQR 0.7–1.3) in the PROCHOB, moderate proteinuric glomerular disease, severe proteinuric glomerular disease, Dent disease, and proteinuria-negative control groups, respectively. Post-hoc Dunn’s test with Bonferroni correction exhibited higher uMB/Cr ratio in the PROCHOB, severe proteinuric glomerular disease, and Dent disease groups than that in the moderate proteinuric glomerular disease group (all adjusted p < 0.01). Additionally, uMB/Cr ratio was lower in the proteinuria-negative controls than that in the PROCHOB, severe proteinuric glomerular disease, and Dent disease groups (all adjusted p < 0.001). Consistent with urinary myoglobin concentrations, uMB/Cr was higher in the PROCHOB group than that in the moderate proteinuric glomerular disease group (Mann–Whitney U test, p < 0.001; Fig. 2C).

In the severe proteinuric glomerular disease group, urinary myoglobin levels tended to reduce as proteinuria improved with treatment (Supplementary Fig. 1). Additionally, urinary myoglobin levels measured from frozen samples stored in non-dedicated containers were significantly lower than those measured from samples collected using dedicated tubes, even within the same patient (Supplementary Fig. 2). Moreover, in two patients, urinary myoglobin levels were measured on the day of kidney biopsy and the following day, and no post-biopsy increase in urinary myoglobin levels was observed, indicating that the biopsy procedure did not affect urinary myoglobin levels.

#### Determination of the Optimal Urinary Myoglobin and uMB/Cr Cutoff Value for PROCHOB Screening

Receiver operating characteristic (ROC) curve analyses comparing patients with PROCHOB and moderate proteinuric glomerular disease were performed using urinary myoglobin concentrations and uMB/Cr ratios. Both markers demonstrated excellent discrimination, with AUCs of 0.995 and 0.998 for myoglobin and uMB/Cr, respectively (Figure 4). Using the optimal cutoff values determined by the Youden index, a myoglobin cutoff of 31.5 ng/mL yielded a sensitivity of 96.0 %, specificity of 100.0 %, positive predictive value of 100.0 %, and negative predictive value of 97.1 %. A uMB/Cr cutoff of 22.2 ng/mgCr yielded a sensitivity of 100 %, specificity of 94.0 %, positive predictive value of 92.6 %, and negative predictive value of 100 %. Because it normalizes urine concentration and prioritizes high sensitivity as a biomarker, uMB/Cr may be more robust for clinical implementation, despite comparable AUCs.

**Figure 4.**
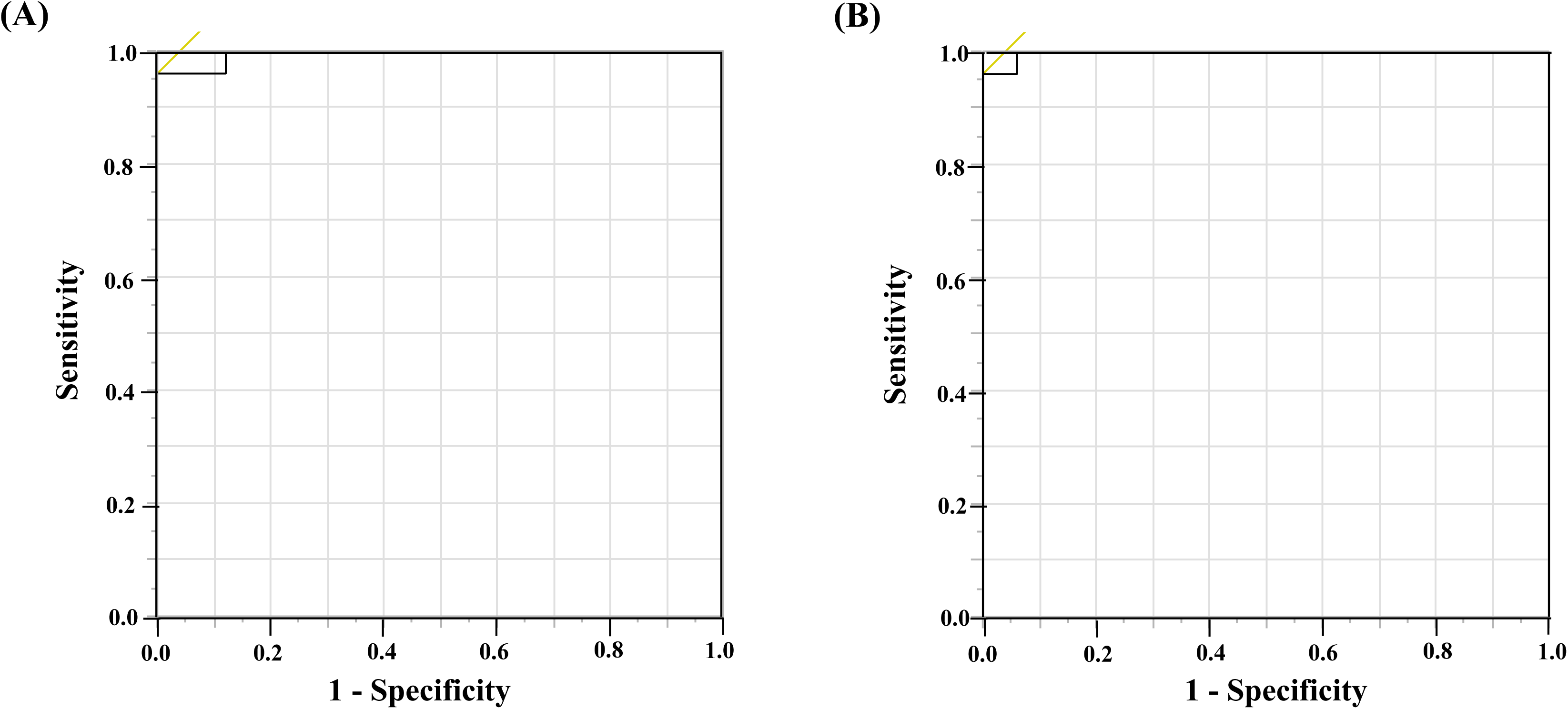
ROC curves for urinary myoglobin and the myoglobin-to-creatinine ratio to discriminate chronic benign proteinuria from moderate proteinuric glomerular diseases. (A) ROC curve for urinary myoglobin. (B) ROC curve for the myoglobin-to-creatinine ratio. AUC is calculated, and optimal cutoff values are determined using the Youden index.

To further differentiate Dent disease from PROCHOB, we assessed the uBMG and uBMG/Cr ratios using ROC analysis. Using the optimal cutoff values determined by the Youden index, uBMG and uBMG/Cr cutoffs of 11,764 μg/L and 15.23 μg/mgCr, respectively, yielded a sensitivity of 100.0 %, specificity of 100.0 %, positive predictive value of 100.0 %, and negative predictive value of 100.0 % for distinguishing Dent disease (n = 6) from PROCHOB (n = 25) (AUC = 1.00 for both; p < 0.0001).

Based on these findings, Figure 5 presents a practical screening algorithm for patients with mild proteinuria and preserved kidney function using the uBMG/Cr and uMB/Cr ratios. For clinical applicability, the ROC-derived optimal cutoffs (15.23 µg/mgCr and 22.2 ng/mgCr for uBMG/Cr and uMB/Cr, respectively) were rounded to convenient integer thresholds (15 µg/mgCr and 22 ng/mgCr, respectively). Using these thresholds, patients were classified into three patterns: (i) uBMG/Cr high and uMB/Cr high (uBMG/Cr ≥ 15 and uMB/Cr ≥ 22)—indicating Dent disease; (ii) uBMG/Cr low and uMB/Cr high (uBMG/Cr < 15 and uMB/Cr ≥ 22)—indicating PROCHOB; and (iii) uBMG/Cr low and uMB/Cr low (uBMG/Cr < 15 and uMB/Cr < 22)—indicating glomerular diseases.

**Figure 5.**
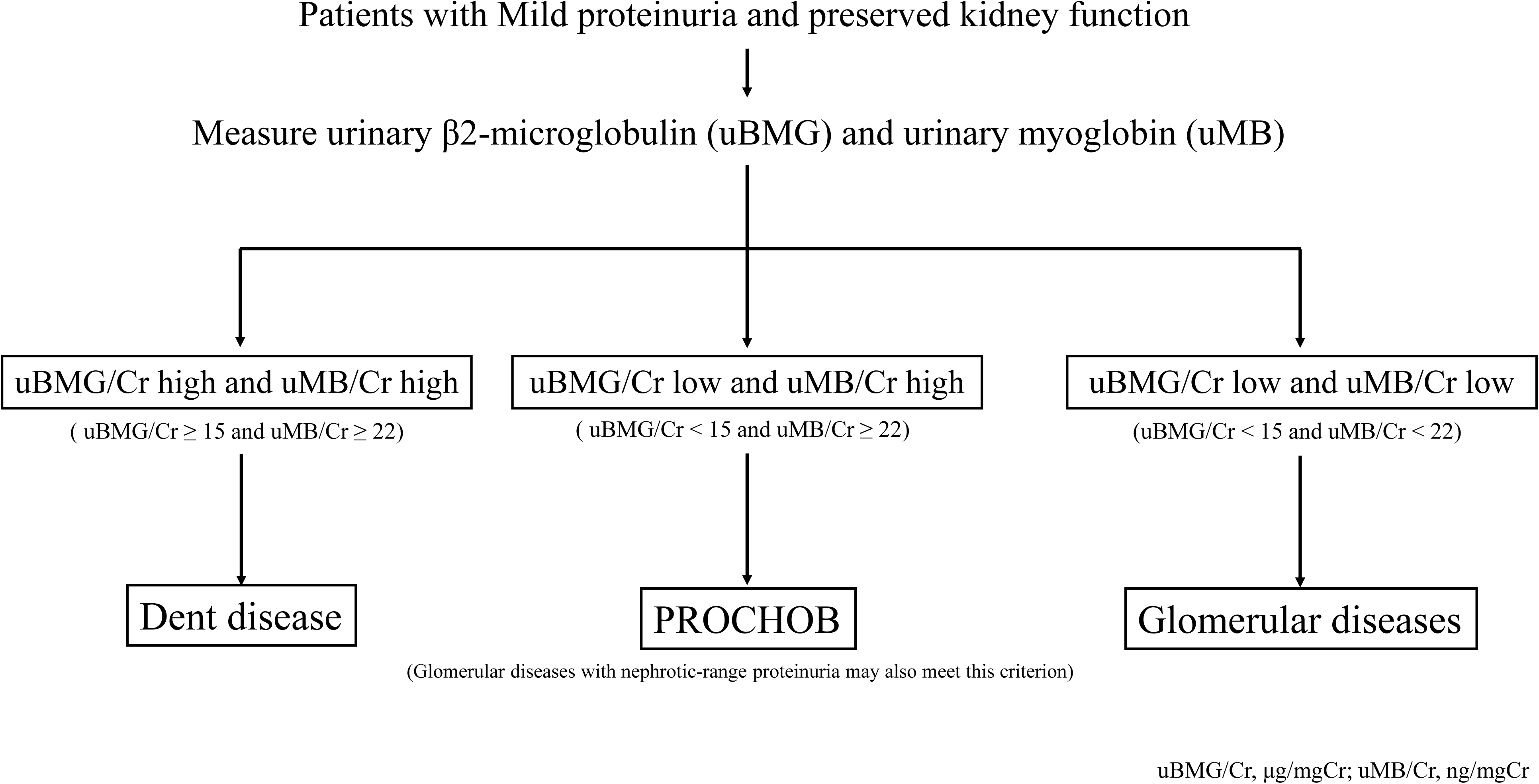
Proposed screening algorithm for suspected chronic benign proteinuria using urinary β2-microglobulin (uBMG) and myoglobin. In patients with mild proteinuria and preserved kidney function, uBMG and urinary myoglobin (uMB) are measured, and the uBMG-to-creatinine ratio (uBMG/Cr) and uMB-to-creatinine ratio (uMB/Cr) are used for stratification. Patients are classified into three categories: (1) uBMG/Cr high and uMB/Cr high (uBMG/Cr ≥ 15 µg/mgCr and uMB/Cr ≥ 22 ng/mgCr)—indicating Dent disease; (2) uBMG/Cr low and uMB/Cr high (uBMG/Cr < 15 µg/mgCr and uMB/Cr ≥ 22 ng/mgCr)—indicating PROCHOB; and (3) uBMG/Cr low and uMB/Cr low (uBMG/Cr < 15 µg/mgCr and uMB/Cr < 22 ng/mgCr)—indicating glomerular diseases.

## Discussion

In this study, we aimed to identify a screening biomarker capable of distinguishing PROCHOB from other glomerular diseases. Proteomic analysis using SomaScan identified myoglobin as a candidate protein. For validation, we expanded the PROCHOB cohort to include 25 patients and a control group with glomerular diseases. Urinary myoglobin levels were measured using a commercially available chemiluminescent enzyme immunoassay (CLEIA)-based assay, and comparative analyses were performed. Based on these findings, we propose a novel diagnostic approach for pediatric patients presenting with moderate proteinuria, in which the uMB/Cr ratio can serve as a screening biomarker. Specifically, when the ratio is ≥ 22.2 ng/mgCr, genetic testing can be performed before a kidney biopsy, enabling a noninvasive diagnosis of PROCHOB.

Myoglobin is a 17 kDa heme protein that is freely filtered through the glomerulus, reabsorbed by proximal tubular epithelial cells, and rapidly cleared from plasma with a half-life of approximately 2–3 hours (1, 2, 24). Megalin and cubilin bind myoglobin and mediate its uptake, with inhibition or genetic deficiency of these receptors in mice resulting in almost complete urinary loss of the injected myoglobin (8). Extending these findings to humans, our in vivo data indicate that impaired receptor-mediated reabsorption because of cubilin dysfunction in PROCHOB can increase urinary myoglobin, serving as a marker of proximal tubular dysfunction. From a practical standpoint, urinary myoglobin is highly unstable and rapidly degrades if not collected in dedicated containers, potentially resulting in falsely low values. Urine myoglobin deteriorates rapidly, with up to 95 % activity loss within 2 hours at 37 °C and a 40 % reduction after 24 hours at -20°C (25). In contrast, it remains relatively stable for up to 3 hours at room temperature and exhibits less deterioration after 24 hours when the urine pH is maintained between 6.5 and 8.8 (25). Consistent with this, we observed lower myoglobin concentrations in frozen specimens stored in non-dedicated containers, even within the same patient. These results indicate that accurate quantification requires prompt collection in dedicated tubes containing an alkalinizing stabilizer and immediate pH adjustment at the time of sampling to minimize degradation.

We observed modest increases in urinary myoglobin levels in the PROCHOB group and in patients with Dent disease and glomerular diseases. In Dent disease, pathogenic variants in *CLCN5* impaired ClC-5 activity and disrupted megalin- and cubilin-mediated receptor endocytosis in the proximal tubules. Accordingly, increased urinary myoglobin levels are biologically plausible and were observed in all Dent disease cases in our cohort. Notably, uBMG was significantly increased only in Dent disease, whereas it remained within the normal range in PROCHOB, consistent with previous studies—indicating that these two tubular disorders are readily distinguishable in clinical practice. In contrast, increased urinary myoglobin in glomerular diseases—specifically in cases of nephrotic-range proteinuria or significant hematuria—may reflect the competitive saturation of megalin/cubilin-mediated proximal tubular uptake by increased intratubular levels of hemoglobin and filtered proteins rather than a primary receptor defect. Because hemoglobin reabsorption depends on megalin/cubilin and myoglobin can partially inhibit hemoglobin uptake, these findings support a shared endocytic pathway [7]. Similarly, albumin and other low-molecular-weight proteins may compete for this receptor system (22). Notably, nephrotic-range proteinuria or significant hematuria is typically not observed in PROCHOB. Therefore, urinary myoglobin can serve as a specific screening marker in patients with mild proteinuria suspected of PROCHOB, similar to uBMG in Dent disease, enabling a genetic-first approach with fewer biopsies.

Urinary myoglobin levels primarily increase in conditions involving significant muscle injury, including post-surgical states, trauma, rhabdomyolysis, drug-induced myopathy, infectious myositis, and acute cardiac conditions, such as myocardial infarction. In severe rhabdomyolysis, urinary myoglobin concentrations can exceed 300,000 μg/L, resulting in dark-colored urine and secondary kidney injury, whereas milder cases may present without visibly discolored urine (24). In healthy individuals, urinary myoglobin is typically undetectable, with previous studies reporting no increase even after strenuous exercise, such as marathon running or a combined 3-hour weight-lifting session followed by a 16-mile run (25). Consistent with these findings, none of the healthy controls in our study exhibited increased levels of urinary myoglobin. In our cohort, urinary myoglobin levels did not increase after kidney biopsy. Therefore, special precautions are generally unnecessary in routine practice, but results should be interpreted cautiously after major trauma or during severe catabolic/consumptive illness.

In this study, SomaScan analysis identified 23 urinary proteins, including myoglobin, that met the pre-specified nomination thresholds in the PROCHOB group. One significant candidate was α-amylase 1 (salivary amylase), previously reported as a megalin ligand. It comprises salivary (AMY1) and pancreatic (AMY2) isoforms and is thought to be predominantly reabsorbed in the proximal tubules via megalin, as supported by receptor-associated protein-deficient mouse experiments and megalin-binding assays using surface plasmon resonance (26). Although no previous studies have directly assessed whether cubilin serves as an amylase ligand, our findings indicate this. However, the direct quantification of these proteins in patients with PROCHOB demonstrated no significant increase, indicating limited biomarker utility. This SomaScan–immunoassay discrepancy may reflect (i) SomaScan’s ability to detect fragmented or modified isoforms that are not captured by conventional immunoassays and (ii) transient fluctuations in urinary excretion that may be missed by single-time-point sampling. Consistent with these findings, we observed no clear increase in the urinary levels of established cubilin-associated proteins reported in animal models and human cubilin-related disorders (3–7, 27–29). This indicates that known cubilin ligands, such as alpha-1-microglobulin have limited usefulness as biomarkers for PROCHOB, potentially because of compensatory proximal tubular uptake and inter-individual variability (11, 14).

This study has several limitations. First, it was conducted in a Japanese cohort with a relatively small sample size because of the rarity of the disease. Therefore, larger multicenter studies and external validation in other ethnic populations are required to confirm the generalizability of our findings. Second, urinary myoglobin is inherently unstable. Although we used a commercially available kit and promptly aliquoted samples to ensure reliable measurements, the possibility of slight underestimation cannot be excluded. Third, the cutoff values incorporated into our proposed screening algorithm (uMB/Cr and uBMG/Cr: 22 ng/mgCr and 15 µg/mgCr, respectively) were derived from a single cohort and specific assay platforms. Therefore, their generalizability to other populations and measurement methods remains unclear. External validation in independent cohorts is warranted, specifically across diverse ethnic and geographic backgrounds, because reference ranges and optimal thresholds may vary by region and ancestry.

This study demonstrated that measuring the uMB/Cr ratio in patients suspected of having PROCHOB can facilitate a noninvasive diagnostic approach before kidney biopsy. Using this biomarker may help avoid unnecessary invasive procedures and treatments, thereby enhancing diagnostic efficiency. Future studies with larger multiethnic cohorts are warranted to confirm these findings.

## Methods

### Sex as a biological variable

This study included male and female participants. Sex distribution and its potential effects on urinary biomarker levels were assessed in all analyses. No significant sex-based differences were observed in the urinary myoglobin concentrations or uMB/Cr ratios. Therefore, the findings are expected to be generalizable to both sexes.

### SomaScan Proteomics

We performed SomaScan-based proteomic analysis of urine samples from seven patients with PROCHOB, six with Alport syndrome, and seven with INS—all genetically diagnosed at Kobe University through targeted panel sequencing (Supplementary Table 6). The patient characteristics are listed in Supplementary Table 1. A 100 μL urine sample from each participant was immediately stored at -80 °C and analyzed using the SomaScan Version 4.1 platform (SomaLogic, Boulder, CO)—that quantifies 7,288 human proteins per sample. The assay was performed following the manufacturer’s standard biofluid protocols (22), including pH adjustment, buffer exchange, and SOMAmer-based hybridization detection, as previously described. Quality control included pooled urine and buffer-only controls, and all samples passed the calibration and normalization criteria of SomaLogic. Statistical analyses of the proteomic data were performed using SomaLogic-provided tools. P values were adjusted for multiple testing across all 7,288 proteins using the Benjamini–Hochberg FDR procedure, and FDR-adjusted q values are reported for transparency. Because the discovery cohort was small, we pre-specified relaxed nomination thresholds (fold change ≥ 2.5 and nominal p < 0.05) to select candidates for orthogonal validation, following previous SomaScan studies (30). For candidate prioritization, we focused on proteins with established biological plausibility as cubilin/megalin-reabsorbed ligands. Accordingly, myoglobin and α-amylase 1 were selected as candidate biomarkers for validation (1, 8, 26). Both analytes are measurable using commercially available assays and were selected as candidate biomarkers for validation.

### Assessment of amylase as a potential biomarker

Urinary amylase activity was quantified in patients with PROCHOB using a commercially available enzyme-based assay (Et-G7-PNP method)—standardized based on JSCC guidelines and analyzed by LSI Medience Corporation (Tokyo, Japan).

### Study Design and Patient Characteristics of the Myoglobin Validation Cohort

The validation cohort included patients with genetically confirmed PROCHOB from Kobe University, including those who underwent SomaScan analysis. Control samples were obtained from our outpatient clinic and selected to represent four groups—(ⅰ) glomerular diseases with moderate proteinuria comparable to PROCHOB, (ⅱ) glomerular diseases with severe proteinuria, (ⅲ) a tubular disorder (Dent disease), and (ⅳ) asymptomatic hematuria or non-kidney diseases without proteinuria. Glomerular disease controls comprised patients with INS, IgA nephropathy, IgA vasculitis nephritis, ANCA-associated vasculitis, hereditary nephrotic syndrome, C3 nephritis, and asymptomatic proteinuria. Moderate proteinuria was defined as an early morning spot urine protein-to-creatinine ratio (uTP/Cr) of 0.15–4.0 g/gCr, and nephrotic-range (severe) proteinuria was defined as uTP/Cr > 4.0 g/gCr. All patients with proteinuria (uTP/Cr > 0.15 g/gCr) were included, except those with asymptomatic hematuria and non-kidney diseases. During urine collection, none of the patients had a history or clinical evidence of conditions known to increase myoglobin production, such as recent vigorous exercise, muscle injury, or rhabdomyolysis.

Patients were stratified into five groups for analysis—the PROCHOB, moderate proteinuric glomerular diseases, severe proteinuric glomerular diseases, Dent disease, and proteinuria-negative control groups. Clinical and laboratory findings, including urinary myoglobin concentrations and uMB/Cr, were compared across these five groups. We performed ROC analyses to determine the optimal cutoff values for urinary myoglobin and uMB/Cr to differentiate PROCHOB from moderate proteinuric glomerular diseases.

Clinical data, including sex, initial symptoms, age at detection, pathological findings, and laboratory data, were extracted from the medical records. Cr-eGFR (mL/min/1.73 m^2^) was calculated using an equation for Japanese patients aged 2–18 years (31) and a different equation for those aged ≥ 19 years (32).

### Urinary Myoglobin Measurement

Owing to the known instability of urinary myoglobin (25), spot urine samples were promptly collected in dedicated urine myoglobin collection tubes (container No. 73; LSI Medience Corporation, Tokyo, Japan) containing sodium metabisulfite, ProClin, and EDTA-3Na, with a required volume of 6 mL. The samples were stored at room temperature, following the manufacturer’s protocol. Urinary myoglobin concentrations were measured using standardized CLEIA performed by LSI Medience Corporation (Tokyo, Japan). The assay had a lower limit of quantification (LLOQ) of 2.0 ng/mL. For statistical analyses, values below the LLOQ were imputed as 1.0 ng/mL (half the LLOQ). Sensitivity analyses using 2.0 ng/mL yielded similar results.

### Statistics

Statistical analyses were performed using JMP version 14.0 (SAS Institute Japan, Tokyo, Japan). Continuous variables are presented as medians with IQRs, and categorical variables are presented as counts and percentages. We performed exploratory comparisons of continuous variables across the five predefined groups using the Kruskal–Wallis test. When the overall test was significant, pairwise post-hoc comparisons were conducted using Dunn’s test with Bonferroni correction. Categorical variables were compared using Fisher’s exact test.

The primary comparison for the most clinically relevant differential diagnosis was between PROCHOB and moderate proteinuric glomerular diseases. Continuous variables were analyzed using the Mann–Whitney U-test. ROC curve analyses were performed to assess the diagnostic performance of urinary myoglobin and uMB/Cr. The AUC was calculated, optimal cutoff values were determined using the Youden index, and sensitivity, specificity, positive predictive value, and negative predictive value were calculated for the study cohort. A two-sided p-value < 0.05 was considered statistically significant.

### Genetic Analysis

Genomic DNA was isolated from the peripheral blood leukocytes of patients using the Quick Gene Mini 80 System or QuickGene-Auto 12S (Kurabo Industries Ltd., Tokyo, Japan), following the manufacturer’s protocol. Targeted panel sequencing was performed for inherited kidney disease-associated genes (Supplementary Table 6). Sequencing samples were prepared using the Haloplex or SureSelect target enrichment system kit (Agilent Technologies, Santa Clara, CA, USA) following the manufacturer’s protocol. Indexed DNA samples were amplified using PCR and sequenced using the MiSeq platform (Illumina, San Diego, CA, USA). Sequence data were processed using two analysis pipelines based on the sequencing date. Reads generated up to May 2024, reads were processed using SureCall software (Agilent Technologies), from alignment through variant calling and categorization. For data generated from June 2024 onward, variant calling was performed according to the Genome Analysis Toolkit (GATK) Best Practices workflows developed by the Broad Institute (33), and variants were annotated using ANNOVAR (34). In patients with suspected PROCHOB and a heterozygous pathogenic variant identified by panel sequencing, we performed custom array comparative genomic hybridization to assess large genomic rearrangements of *CUBN*.

Pathogenicity was assessed based on the ACMG guidelines (35). Candidate exonic variants were assessed using Sorting Intolerant From Tolerant (https://sift.bii.a-star.edu.sg/), PolyPhen-2 (https://genetics.bwh.harvard.edu/pph2/), MutationTaster (https://www.mutationtaster.org/), and Combined Annotation Dependent Depletion (https://cadd.gs.washington.edu/snv). Variants were classified as likely pathogenic based on the criteria described in a previous study of patients with PROCHOB (11) when ≥ 2 tools predicted deleteriousness. Splicing variants in the coding regions or essential splice sites were assessed using a minigene assay (36). Previously reported variants were identified by cross-referencing the Human Gene Mutation Database.

### Study approval

This study was approved by the Institutional Review Board of Kobe University Graduate School of Medicine (IRB No. 301, B230224) and adhered to the ethical guidelines of the Japanese Ministry of Health, Labor and Welfare, and the Declaration of Helsinki. Written informed consent was obtained from all patients and/or their parents before genetic testing. For biomarker measurements, patients were informed using an opt-out approach, and verbal consent was obtained.

## Data availability

All data supporting the findings of this study are available from the corresponding author upon reasonable request. SomaScan proteomic data, processed urinary myoglobin measurements, and associated clinical metadata will be shared in a de-identified form for non-commercial academic purposes.

## Disclosures

A patent application has been submitted covering aspects of the work described in this manuscript.

## Author contributions

YI conducted the study, contributed to the study design, collected data, supervised the analysis, and drafted the manuscript. NS, SI, AY, S A, YK, YI, YT, AK, TY, SI, YA, TA, JF, SF, RH, NI, HK, KK, AK, YK, NK, HM, YO, SS, RS, MW, YY, YY, KI, and CN reviewed and edited the manuscript. THand KN supervised the study and revised the manuscript. All authors have read and approved the final manuscript.

## Funding

Kandai Nozu receives the Grants-in-Aid for Scientific Research (KAKENHI, 23K07698), Childhood-onset, rare and intractable kidney diseases in Japan, Research on rare and intractable diseases, Health, Labour and Welfare Sciences Research Grants (23FC1047), the Japan Agency for Medical Research and Development (AMED) (Grant Number 24015773 and 22810094 and 23ek0109617s1702)

## Acknowledgments

We acknowledge SomaLogic, Inc. (Boulder, CO, USA) and FonesLife Corporation for their support in conducting the SomaScan proteomic measurements.

## Data sharing statement

The data underlying this article will be shared upon reasonable request by the corresponding Authors.

## Notes

**Conflict of Interest:** The authors have declared that no conflict of interest exists.

### Competing Interest Statement

The authors have declared no competing interest.

### Author Declarations

This study was approved by the Institutional Review Board of Kobe University Graduate School of Medicine (IRB No. 301, B230224) and adhered to the ethical guidelines of the Japanese Ministry of Health, Labor and Welfare, and the Declaration of Helsinki.

