## Supplemental Files for "Proteomic Discovery of Urinary Myoglobin as a Noninvasive Biomarker for PROCHOB caused by *CUBN* Variants"

Tomoko Horinouchi, MD, PhD

Department of Pediatrics, Kobe University Graduate School of Medicine

7-5-1 Kusunoki-cho, Chuo, Kobe, Hyogo 650-0017, Japan

ORCID: https://orcid.org/0000-0003-1655-6030

### Supplementary Table 1. Patient characteristics of individuals who underwent SomaScan analysis

| Patinet ID | Diagnosis | Age at first detection of proteinuria (year) | Age at analysiss (year) | Gender | Cr-eGFR (mL/min/1.73 m²) | S-Alb (g/dL) | uPro/Cr (g/gCr) | uBMG (μg/L) | Pathology | Gene | Variant 1 | | Variant 2 | |
|---|---|---|---|---|---|---|---|---|---|---|---|---|---|---|
| | | | | | | | | | | | Nucleotide change | Amino Acid change | Nucleotide change | Amino Acid change |
| Neph558 | PROCHOB | 1–5 | 6–10 | F | 110.5 | 4.5 | 1.5 | 96 | MGA | *CUBN* | c.6821+3A>G | – | c.7580_7581del | p. (Cys2527*) |
| Neph558-1 | PROCHOB | 1–5 | 11–15 | M | 96.0 | 4.4 | 0.78 | 99 | MGA | *CUBN* | c.6821+3A>G | – | c.7580_7581del | p. (Cys2527*) |
| Neph697 | PROCHOB | 1–5 | 6–10 | M | 98.1 | 4.4 | 1.28 | 310 | MGA | *CUBN* | c.6821+3A>G | – | c.5519dupG | p. (Thr1841Hisfs*14) |
| Neph697-1 | PROCHOB | 1–5 | 6–10 | F | 123.8 | 4.6 | 0.79 | 199 | MGA | *CUBN* | c.6821+3A>G | – | c.5519dupG | p. (Thr1841Hisfs*14) |
| Neph711 | PROCHOB | 1–5 | 16–20 | F | 126.8 | 4.1 | 0.46 | 129 | MGA | *CUBN* | c.5291G>A | p. (Cys1764Tyr) | c.5842G>A | p. (Asp1948Asn) |
| Neph716 | PROCHOB | 1–5 | 11–15 | M | 115.1 | 4.5 | 0.7 | 72 | MGA | *CUBN* | c.7432T>C | p. (Cys2478Arg) | c.10562C>G | p. (Ser3521*) |
| Neph719 | PROCHOB | 1–5 | 6–10 | M | 107.7 | 4.5 | 0.51 | 200 | MGA | *CUBN* | c.5291G>A | p. (Cys1764Tyr) | c.5806_5807delinsAA | p. (Ser1936Asn) |

| | | | | | | | | | | | | | | |
|---|---|---|---|---|---|---|---|---|---|---|---|---|---|---|
| A743 | AS | 1–5 | 11–15 | M | 126 | 3.5 | 0.57 | 174 | – | *COL4A5* | c.3907G>T | p. (Gly1303*) | – | – |
| A1103 | AS | 1–5 | 1–5 | M | 108.9 | 4.2 | 0.29 | 86 | MGA | *COL4A5* | c.1914_1940del | p. (Val639_647del) | – | – |
| A1267 | AS | 1–5 | 1–5 | F | 170 | 4 | 0.83 | 105 | MGA | *COL4A3* | Exons 18–19 deletion | – | COL4A3–COL4A4 12,413-bp deletion | – |
| A1285 | AS | 1–5 | 11–15 | M | 80.1 | 3.5 | 0.6 | 56 | – | *COL4A5* | c.1543G>A | p. (Gly515Arg) | – | – |
| A1285-1 | AS | 1–5 | 11–15 | M | 51.2 | 3.9 | 0.42 | 61 | – | *COL4A5* | c.1543G>A | p. (Gly515Arg) | – | – |
| A1353 | AS | 1–5 | 25–30 | F | 72.3 | 4.2 | 1.1 | 93 | MGA | *COL4A5* | c.5029C>T | p. (Arg1677*) | – | – |
| INS-1 | INS | 6–10 | 6–10 | M | 114.2 | 2.9 | 2.9 | 141 | MGA | – | – | – | – | – |
| INS-2 | INS | 11–15 | 11–15 | M | 135.5 | 3.2 | 4.66 | 198 | – | – | – | – | – | – |
| INS-3 | INS | 1–5 | 1–5 | M | 127.5 | 1.4 | 6.74 | 125 | MGA | – | – | – | – | – |
| INS-4 | INS | 6–10 | 6–10 | F | 118.9 | 1.5 | 26.22 | 396 | – | – | – | – | – | – |
| INS-5 | INS | 11–15 | 11–15 | M | 119.1 | 1.9 | 19.76 | 346 | FSGS | Not detected | – | – | – | – |
| INS-6 | INS | 1–5 | 1–5 | F | 150.3 | 3.4 | 15.03 | 105 | MGA | – | – | – | – | – |
| INS-7 | INS | 1–5 | 6–10 | M | 101.6 | 3.4 | 8.35 | 90 | MGA | – | – | – | – | – |

PROCHOB, chronic benign proteinuria (proteinuria due to biallelic *CUBN* variants); AS, Alport syndrome; INS, idiopathic nephrotic syndrome; Cr-eGFR, creatinine-based estimated glomerular filtration rate; S-Alb, serum albumin; uPro/Cr, urinary protein-to-creatinine ratio; uBMG, urinary β2-microglobulin; MGA, minor glomerular abnormalities; FSGS, focal segmental glomerulosclerosis; F, female; M, male; –, not available/not assessed.

**Supplementary Table 2. Urinary amylase levels in patients with PROCHOB**

| | AMY (reference range 65–840) | s-Amy, % (reference range 17.7–61.3) |
|---|---|---|
| Neph576 | 578 | 65.9 |
| Neph697 | 426 | 54.4 |
| Neph711 | 1936 | 71.0 |
| Neph716 | 17 | 50.3 |
| neph719 | 84 | 83.9 |
| Neph801-1 | 377 | 68.0 |
| Neph913 | 421 | 71.1 |

PROCHOB, chronic benign proteinuria (proteinuria due to biallelic *CUBN* variants); AMY, urinary amylase; s-Amy, salivary-type amylase fraction.

**Supplementary Table 3. Cubilin-Dependent Reabsorbed Proteins in Urine: SomaScan-Based Comparison Between PROCHOB and Other Disease Groups**

| Sequence ID | Protein Name | UniProt ID | Gene Symbol | Fold.Change * | t.statistic | p.value |
|---|---|---|---|---|---|---|
| 17672-184 | Gastric intrinsic factor | P27352 | CBLIF | -0.095 | -0.09 | 0.93 |
| 15589-1 | Vitamin D-binding protein | P02774 | GC | -1.4 | 2.52 | 0.022 |
| 18380-78 | Serum albumin | P02768 | ALB | 0.17 | 0.08 | 0.94 |
| 4915-64 | Hemoglobin | P69905\|P68871 | HBA1\|HBB | 0.28 | -2.55 | 0.025 |
| 4162-54 | Serotransferrin | P02787 | TF | 0.1 | -0.2 | 0.85 |
| 11510-31 | Apolipoprotein L1 | O14791 | APOL1 | -0.0089 | -1.01 | 0.33 |
| 15453-3 | Alpha-1-microglobulin | P02760 | AMBP | 0.0074 | 0.97 | 0.35 |
| 10618-190 | Low-density lipoprotein receptor-related protein 2 (Megalin) | P98164 | LRP2 | 0.77 | -1.51 | 0.16 |
| 4322-28 | Protein amnionless | Q9BXJ7 | AMN | 0.029 | -0.37 | 0.72 |
| 3640-14 | alpha-2-macroglobulin receptor-associated protein (Receptor associated protein) | P30533 | LRPAP1 | -0.3 | 0.3 | 0.77 |
| 3184-25 | Coagulation factor VII (Recombinant activated factor VIIa) | P08709 | F7 | 0.37 | -1.1 | 0.28 |

PROCHOB, chronic benign proteinuria (proteinuria due to biallelic *CUBN* variants). ∗, A negative fold change indicates that the protein is more abundant in the PROCHOB group

### Supplementary Table 4. *CUBN* variant information

| Exon | Nucleotide change | Amino Acid change | Type | dbSNP ID | gnomAD | Tommo54K | *In silico* algorithm predictions for missense variant | | | | ACMG | Reference |
|---|---|---|---|---|---|---|---|---|---|---|---|---|
| | | | | | | | CADD | SIFT | PolyPhen-2 | Mutation Taster | | |
| 26 | c.3685_3686del | p. (Pro1229Lysfs*2) | frameshift | – | Absent | Absent | – | – | – | – | Pathogenic (PVS1+PM2) | – |
| 31 | c.4612T>A | p.(Trp1538Arg) | missense | – | Absent | Absent | 25.6 | Deleterious | probably damaging | Deleterious - | Likely Pathogenic (PM1+PM2+PM3+PP3) | |
| IVS32 | c.4855+2C>G | – | splicing | rs772316719 | 4.10E-06 | 0.000295 | – | – | – | – | Pathogenic (PVS1+PM2+PP5) | (1) |
| 36 | c.5291G>A | p.(Cys1764Tyr) | missense | rs986680465 | 5.47E-06 | 0.000175 | 24.9 | Deleterious | probably damaging | Deleterious | Likely Pathogenic (PM1+PM2+PM3+PP3) | (2) |
| 36 | c.5302_5304del | p.(Ile1768del) | in-frame | rs775161946 | 6.20E-06 | 0.000037 | – | – | – | – | Likely Pathogenic (PM2+PM3+PM4) | – |
| 37 | c.5519dupG | p.(Thr1841Hisfs*14) | frameshift | - | Absent | Absent | – | – | – | – | Pathogenic (PVS1+PM2) | – |
| 39 | c.5806_5807delinsAA | p.(Ser1936Asn) | missense | rs1554796668 | Absent | Absent | – | Deleterious | probably damaging | – | Likely Pathogenic (PM1+PM2+PM3) | – |
| 39 | c.5842G>A | p.(Asp1948Asn) | missense | rs1285741474 | 7.44E-06 | 0.000009 | 24.4 | Tolerated | probably damaging | benign | Likely Pathogenic (PM1+PM2+PM3) | |
| IVS40 | c.6125-2A>G | – | splicing | rs75386064 | 0.000460 | Absent | – | – | – | – | Likely Pathogenic (PVS1+PP5) | (3) |
| IVS44 | c.6821+3A>G | – | splicing | rs767078847 | 1.613E-05 | 0.000203 | – | – | – | – | Likely Pathogenic (PS3+PM2) | (4) |
| 48 | c.7352-3_7354 del | – | splicing | rs1841809257 | 0 | 0.000018 | – | – | – | – | Pathogenic (PVS1+PM2) | – |
| 48 | c.7432T>C | p.(Cys2478Arg) | missense | – | Absent | Absent | 27.1 | Deleterious | probably damaging | benign | Likely Pathogenic (PM1+PM2+PM3+PP3+BP4) | – |
| 49 | c.7543_7544delinsAA | p.(Gly2515Asn) | missense | – | Absent | Absent | – | Deleterious | probably damaging | – | Likely Pathogenic (PM1, PM2, PM3) | – |

| | | | | | | | | | | | | |
|---|---|---|---|---|---|---|---|---|---|---|---|---|
| 49 | c.7580_7581del | p.(Cys2527*) | nonsense | – | 3.19E-06 | 0.000028 | – | – | – | – | Pathogenic (PVS1+PM2) | – |
| 49-50 | heterozygous deletion in exon 49-50 | – | – | – | – | – | – | – | – | – | Pathogenic (PVS1+PM2) | – |
| 52 | c.8080A>G | p.(Ile2694Val) | missense | rs773503032 | Absent | Absent | 17.8 | Tolerated | benign | benign | Uncertain Significance (PM1+PM2+PM3+BP4) | – |
| IVS52 | c.8184+1G>T | – | splicing | – | Absent | Absent | – | – | – | – | Pathogenic (PVS1+PM2) | – |
| IVS58 | c.9236+5G>A | – | splicing | rs1458087960 | 8.05E-06 | 0.000013 | – | – | – | – | Pathogenic (PVS1+PM2) | – |
| IVS58 | c.9237-13_9237-10del | – | splicing | – | 2.80E-05 | Absent | – | – | – | – | Likely Pathogenic (PS3+PM2) | – |
| 63 | c.10055_10058del | p. (Gln3352Pfofs*16) | frameshift | – | Absent | Absent | – | – | – | – | Pathogenic (PVS1+PM2) | – |
| 64 | c.10245C>A | p.(Tyr3415*) | nonsense | rs147730705 | 2.48E-06 | 0.000203 | – | – | – | – | Pathogenic (PVS1+PM2+PP5) | (5) |
| 64 | c.10342T>G | p.(Cys3448Gly) | missense | – | Absent | Absent | 25.0 | Tolerated | probably damaging | Deleterious | Likely Pathogenic (PM1+PM2+PM3+PP3) | – |
| 66 | c.10535G>A | p.(Gly3512Asp) | missense | -– | Absent | Absent | 25.1 | Deleterious | probably damaging | Deleterious | Likely Pathogenic (PM1+PM2+PM3+PP3) | – |
| 66 | c.10562C>G | p.(Ser3521*) | nonsense | - | Absent | Absent | – | – | – | – | Pathogenic (PVS1+PM2) | – |
| 54-55 | heterozygous deletion in exon 54-55 | – | – | – | – | – | – | – | – | – | Pathogenic (PVS1+PM2) | – |

### Supplementary Table 5. Clinical findings in the validation cohort

| Patient ID | Diagnosis | Age at detection (years) | Age at laboratory testing (years) | Sex | Cr-eGFR (mL/min/1.73 m²) | sAlb (g/dL) | uMB (ng/mL) | uMB/Cr (ng/mgCr) | uB MG(μg/L) | uCr (mg/dL) | Urinary occult blood | uTP/Cr (g/gCr) | uBMG/cr (μg/mg Cr) | Pathologic findings | Gene | Variant 1 | Variant 2 |
|---|---|---|---|---|---|---|---|---|---|---|---|---|---|---|---|---|---|
| Neph255 | PROCHOB | 1–5 | 6–10 | M | 131.1 | 4.5 | 250 | 204.08 | 257 | 122.5 | - | 0.76 | 0.21 | MCD | *CUBN* | c.5291G>A | c.5842G>A |
| Neph255-1 | PROCHOB | 1–5 | 6–10 | F | 141.9 | 3.7 | 365 | 323.29 | 267 | 112.9 | - | 0.83 | 0.24 | – | *CUBN* | c.5291G>A | c.5842G>A |
| Neph558 | PROCHOB | 1–5 | 6–10 | F | 110.5 | 4.4 | 168 | 691.07 | 96 | 24.31 | - | 1.5 | 0.39 | MCD | *CUBN* | c.6821+3A>G | c.7580_7581del |
| Neph558-1 | PROCHOB | 1–5 | 11–15 | M | 96 | 4.5 | 49.8 | 62.64 | 99 | 79.5 | - | 0.89 | 0.12 | MCD | *CUBN* | c.6821+3A>G | c.7580_7581del |
| Neph576 | PROCHOB | 1–5 | 21–25 | M | 105 | 5 | 265 | 115.72 | 214 | 229 | - | 0.55 | 0.09 | MCD | *CUBN* | c.4855+2C>G | c.7352-3_7354del |
| Neph580 | PROCHOB | 1–5 | 11–15 | F | 157.7 | 4.7 | 112 | 125.84 | 104.3 | 89 | - | 0.58 | 0.12 | MCD | *CUBN* | c.1811C>T | c.5854T>C |
| Neph642 | PROCHOB | 1–5 | 6–10 | M | 141 | 4.5 | 31.5 | 36.37 | 26 | 86.6 | - | 1 | 0.03 | MCD | *CUBN* | c.8080A>G | c.9236+5G>A |
| Neph697 | PROCHOB | 1–5 | 6–10 | M | 90.4 | 4.4 | 554.6 | 335.71 | 394 | 165.2 | - | 0.79 | 0.24 | MCD | *CUBN* | c.5519dupG | c.6821+3A>G |
| Neph697-1 | PROCHOB | 1–5 | 11–15 | F | 113.7 | 4.6 | 506 | 793.73 | 224 | 63.75 | - | 0.89 | 0.35 | MCD | *CUBN* | c.5519dupG | c.6821+3A>G |
| Neph711 | PROCHOB | 1–5 | 16–20 | F | 126.8 | 4.1 | 1010 | 574.84 | 294 | 175.7 | - | 0.46 | 0.17 | MCD | *CUBN* | c.5291G>A | c.5842G>A |
| Neph749 | PROCHOB | 1–5 | 1–5 | F | 108.8 | 4.7 | 44.1 | 72.93 | 190 | 60.47 | - | 0.61 | 0.31 | – | *CUBN* | c.5302_5304delATC | c.10535G>A |
| Neph800 | PROCHOB | 1–5 | 6–10 | M | 114 | 4.1 | 67.1 | 124.95 | 118 | 53.7 | - | 0.89 | 0.22 | MCD | *CUBN* | c.3685_3686delCC | c.6125-2A>G |
| Neph800-1 | PROCHOB | 1–5 | 1–5 | F | 116 | 4.6 | 69.9 | 145.32 | 136 | 48.1 | - | 1.49 | 0.28 | – | *CUBN* | c.3685_3686delCC | c.6125-2A>G |
| Neph819 | PROCHOB | 1–5 | 6–10 | M | 126 | 4.2 | 10.9 | 22.20 | 167 | 49.1 | + | 0.55 | 0.34 | MCD | *CUBN* | c.10245C>A | c.5806_5807delinsAA |
| Neph845 | PROCHOB | 6–10 | 11–15 | F | 137 | 4.1 | 64.7 | 131.24 | 133 | 49.3 | - | 0.43 | 0.27 | MCD | *CUBN* | c.5291G>A | c.5302_5304del |
| Neph913 | PROCHOB | 1–5 | 6–10 | M | 137.6 | 4 | 193 | 414.16 | 87 | 46.6 | - | 0.62 | 0.19 | – | *CUBN* | c.5291G>A | c.7543_7544delinsAA |
| Neph945 | PROCHOB | 1–5 | 1–5 | F | 102.3 | 4 | 254 | 260.51 | 650 | 97.5 | - | 1.2 | 0.67 | – | *CUBN* | c.8184+1G>T | c.9237-13_9237-10del |
| Neph946 | PROCHOB | 1–5 | 1–5 | F | 137.5 | 4.3 | 79.3 | 299.25 | 145 | 26.5 | + | 1.58 | 0.55 | MCD | *CUBN* | c.4855+2C>G | c.10055_10058del |

| | | | | | | | | | | | | | | | | | |
|---|---|---|---|---|---|---|---|---|---|---|---|---|---|---|---|---|---|
| Neph956 | PROCHOB | 1–5 | 11–15 | F | 112.4 | 4.9 | 219 | 229.56 | 127 | 95.4 | - | 0.75 | 0.13 | MCD | *CUBN* | c.4855+2C>G | c.5291G>A |
| Neph985 | PROCHOB | 6–10 | 26–30 | M | 84 | 4.2 | 997 | 1508.32 | 282 | 66.1 | - | 0.49 | 0.43 | MCD | *CUBN* | c.7580_7581del | Large deletion (intron48-50) |
| Neph1020 | PROCHOB | 1–5 | 11–15 | M | 142 | 4.3 | 326 | 493.19 | 78 | 66.1 | - | 0.62 | 0.12 | MCD | *CUBN* | c.4612T>A | c.6821+3A>G |
| Neph1051 | PROCHOB | 6–10 | 11–15 | M | 90 | 4.7 | 1620 | 470.93 | 143 | 344 | + | 0.38 | 0.04 | – | *CUBN* | c.4855+2C>G | Large deletion (intron54-55) |
| Neph1051-1 | PROCHOB | 1–5 | 11–15 | M | 119 | 4.5 | 204 | 416.33 | 165 | 49 | + | 0.45 | 0.34 | – | *CUBN* | c.4855+2C>G | Large deletion (intron54-55) |
| Neph1051-2 | PROCHOB | 6–10 | 6–10 | M | 90 | 4.5 | 318 | 324.49 | 245 | 98 | + | 0.56 | 0.25 | – | *CUBN* | c.4855+2C>G | Large deletion (intron54-55) |
| A729 | PROCHOB | 1–5 | 16–20 | M | 76 | 4.4 | 124 | 55.96 | 68 | 221.6 | + | 0.32 | 0.03 | MCD | *CUBN* | c.10342T>G | c.10562C>G |
| AS-1 | Alport syndrome | 1–5 | 1–5 | M | 170 | 4 | 1 | 3.80 | 105 | 26.3 | + | 0.37 | 0.40 | MCD | *COL4A3* | Exon18-19 deletion | COL4A3-COL4A4 12413 base deletion |
| AS-2 | Alport syndrome | 1–5 | 6–10 | M | 136 | 4.4 | 2.6 | 2.93 | 79 | 88.6 | + | 0.38 | 0.09 | MCD | *COL4A5* | exon14-17 deletion | – |
| AS-3 | Alport syndrome | 1–5 | 11–15 | M | 167 | 4 | 2.4 | 2.35 | 92 | 102 | + | 0.25 | 0.09 | MCD | *COL4A5* | exon14-19 deletion | – |
| ANCA-1 | ANCA-associated vasculitis | 6–10 | 11–15 | F | 55 | 3.3 | 1 | 3.88 | 219 | 25.8 | + | 1.45 | 0.85 | MCD | Not performed | – | – |
| C3-1 | C3 nephritis | 6–10 | 11–15 | M | 170 | 3.2 | 4.8 | 2.88 | 185 | 166.4 | + | 1.39 | 0.11 | DMP | Not performed | – | – |
| Neph986 | hereditary NS | 1–5 | 1–5 | M | 105.1 | 4.5 | 10.1 | 10.86 | 307 | 93 | - | 1.3 | 0.33 | – | *COQ8B* | c.737G>A | c.737G>A |
| Neph4 | hereditary NS | 1–5 | 11–15 | F | 123.4 | 4.2 | 2.2 | 2.52 | 123 | 87.4 | + | 0.46 | 0.14 | DMP | *LAMB2* | c.225delC | c.2095G>C |
| Neph345 | hereditary NS | 1–5 | 6–10 | F | 232.7 | 0.9 | 3.8 | 6.41 | 174 | 59.3 | - | 4.8 | 0.29 | MCD | *NPHS1* | c.2207T>C | c.3166+5G>A |

| | | | | | | | | | | | | | | | | | |
|---|---|---|---|---|---|---|---|---|---|---|---|---|---|---|---|---|---|
| INS-1 | Idiopathic NS | 1–5 | 1–5 | F | 143.8 | 2.8 | 1 | 2.09 | 73 | 47.9 | - | 0.53 | 0.15 | – | Not performed | – | – |
| INS-2 | Idiopathic NS | 1–5 | 1–5 | F | 140 | 2.8 | 1 | 1.21 | 239 | 82.7 | + | 0.73 | 0.29 | DMP | Not detected | – | – |
| INS-3 | Idiopathic NS | 6–10 | 11–15 | M | 123.3 | 3.5 | 1 | 0.52 | 50 | 193 | - | 1.44 | 0.03 | – | Not performed | – | – |
| INS-4 | Idiopathic NS | 11–15 | 11–15 | F | 148.6 | 3.6 | 4.3 | 7.72 | 170 | 55.7 | - | 1.13 | 0.31 | – | Not performed | – | – |
| INS-5 | Idiopathic NS | 1–5 | 1–5 | M | 144.2 | 3.3 | 16.4 | 23.67 | 73 | 69.3 | - | 15.5 | 0.11 | MCD | Not performed | – | – |
| INS-6 | Idiopathic NS | 11-15 | 11–15 | F | 159.4 | 3.5 | 55.1 | 36.73 | 193 | 150 | - | 6.93 | 0.13 | – | Not performed | – | – |
| INS-7 | Idiopathic NS | 11-15 | 11–15 | F | 101.1 | 2.5 | 202.5 | 45.79 | 299 | 442.2 | - | 4.43 | 0.07 | FSGS | Not detected | – | – |
| INS-8 | Idiopathic NS | 11-15 | 16–20 | F | 110.8 | 2.3 | 140.9 | 73.39 | 237 | 192 | - | 6.95 | 0.12 | – | Not performed | – | – |
| INS-9 | Idiopathic NS | 6–10 | 1–5 | F | 125 | 3.6 | 161.5 | 180.25 | 338 | 89.6 | - | 19.64 | 0.38 | – | Not performed | – | – |
| INS-10 | Idiopathic NS | 11-15 | 11–15 | F | 104.7 | 2.7 | 308 | 643.01 | 98 | 47.9 | - | 17.5 | 0.20 | – | Not performed | – | – |
| INS-11 | Idiopathic NS | 1–5 | 1–5 | F | 111.2 | 0.9 | 78.7 | 90.88 | 2045 | 86.6 | + | 31.1 | 2.36 | – | Not performed | – | – |
| OP-1 | Orthostatic proteinuria | 1–5 | 11–15 | F | 133.3 | 4.8 | 1 | 0.25 | 379 | 392.3 | - | 0.19 | 0.10 | – | Not performed | – | – |
| OP-2 | Orthostatic proteinuria | 11-15 | 11–15 | F | 118.8 | 4.3 | 1 | 0.64 | 135 | 156.7 | - | 0.6 | 0.09 | – | Not performed | – | – |

| | | | | | | | | | | | | | | | | | |
|---|---|---|---|---|---|---|---|---|---|---|---|---|---|---|---|---|---|
| OP-3 | Orthostatic proteinuria | 6–10 | 11–15 | F | 135 | 4.5 | 1 | 0.54 | 277 | 184.2 | - | 0.32 | 0.15 | – | Not detected | – | – |
| OP-4 | Orthostatic proteinuria | 6–10 | 11–15 | M | 119.8 | 4.1 | 1 | 1.87 | 60 | 53.5 | - | 0.42 | 0.11 | – | Not performed | – | – |
| OP-5 | Orthostatic proteinuria | 11-15 | 1–5 | M | 118.4 | 4.3 | 1 | 2.07 | 106 | 48.4 | - | 0.14 | 0.22 | – | Not performed | – | – |
| Neph738 | asymptomatic proteinuria | 1–5 | 1–5 | F | 125.7 | 3.5 | 1 | 0.98 | 59 | 102 | - | 1.13 | 0.06 | FSGS | Not detected | – | – |
| Neph922 | asymptomatic proteinuria | 1–5 | 6–10 | M | 140 | 3.4 | 1 | 2.16 | 114 | 46.2 | + | 0.823 | 0.25 | MCD | Not detected | – | – |
| Neph981 | asymptomatic proteinuria | 1–5 | 1–5 | F | 140.1 | 3.6 | 14.1 | 14.10 | 142 | 100 | + | 1.81 | 0.14 | MCD | Not detected | – | – |
| IgA-VN-1 | IgA vasculitis nephritis | 6–10 | 6–10 | F | 106.1 | 3.6 | 1 | 0.80 | 218 | 125.1 | + | 0.57 | 0.17 | MCD | Not performed | – | – |
| IgA-VN-2 | IgA vasculitis nephritis | 1–5 | 1–5 | M | 106 | 4.1 | 2.7 | 8.11 | 122 | 33.3 | + | 0.99 | 0.37 | MCD | Not performed | – | – |
| IgA-VN-3 | IgA vasculitis nephritis | 11-15 | 11–15 | M | 126 | 3.2 | 8.7 | 9.04 | 151 | 96.2 | + | 1.17 | 0.16 | DMP | Not detected | – | – |
| IgA-VN-4 | IgA vasculitis nephritis | 6–10 | 6–10 | M | 107.6 | 4.1 | 2.2 | 1.63 | 187 | 134.7 | + | 0.22 | 0.14 | – | Not performed | – | – |
| IgA-VN-5 | IgA vasculitis nephritis | 6–10 | 11–15 | F | 107.1 | 4.4 | 3.5 | 1.26 | 239 | 278.1 | + | 0.38 | 0.09 | – | Not detected | – | – |
| IgA-VN-6 | IgA vasculitis nephritis | 6–10 | 6–10 | F | 101.5 | 3.2 | 19.6 | 22.76 | 91 | 86.1 | + | 2.4 | 0.11 | – | Not performed | – | – |
| IgA-VN-7 | IgA vasculitis nephritis | 1–5 | 1–5 | F | 150.4 | 4 | 22.5 | 23.49 | 236 | 95.8 | + | 2.76 | 0.25 | – | Not performed | – | – |

| | | | | | | | | | | | | | | | | | |
|---|---|---|---|---|---|---|---|---|---|---|---|---|---|---|---|---|---|
| IgA-VN-8 | IgA vasculitis nephritis | 11–15 | 11–15 | M | 92.8 | 2.8 | 10.4 | 7.38 | 193 | 140.9 | + | 1.66 | 0.14 | – | Not performed | – | – |
| IgA-VN-9 | IgA vasculitis nephritis | 6–10 | 6–10 | M | 134.5 | 2.7 | 5.3 | 18.09 | 126 | 29.3 | + | 5.03 | 0.43 | MCD | Not performed | – | – |
| IgA-VN-10 | IgA vasculitis nephritis | 6–10 | 11–15 | M | 123 | 2.6 | 31.3 | 45.63 | 236 | 68.6 | + | 5.18 | 0.34 | MCD | Not performed | – | – |
| IgA-VN-11 | IgA vasculitis nephritis | 11–15 | 11–15 | F | 118.3 | 2.9 | 111.9 | 71.27 | 1114 | 157 | + | 5 | 0.71 | MCD | Not performed | – | – |
| IgAN-1 | IgA nephropathy | 6–10 | 11–15 | F | 122.5 | 3.9 | 1 | 0.93 | 91 | 107.6 | + | 0.18 | 0.08 | MCD | Not performed | – | – |
| IgAN-2 | IgA nephropathy | 1–5 | 11–15 | M | 131 | 4.1 | 1 | 1.26 | 116 | 79.5 | + | 0.25 | 0.15 | MCD | Not detected | – | – |
| IgAN-3 | IgA nephropathy | 11–15 | 11–15 | M | 101.8 | 4.5 | 1 | 0.43 | 139 | 233 | + | 0.23 | 0.06 | MCD | Not performed | – | – |
| IgAN-4 | IgA nephropathy | 11–15 | 11–15 | F | 96 | 4.5 | 2.6 | 2.57 | 62 | 101.1 | + | 0.26 | 0.06 | MCD | Not performed | – | – |
| IgAN-5 | IgA nephropathy | 11–15 | 11–15 | M | 83 | 4 | 4.4 | 2.65 | 52 | 166.2 | + | 0.25 | 0.03 | MCD | Not performed | – | – |
| IgAN-6 | IgA nephropathy | 1–5 | 1–5 | M | 110 | 2.9 | 5.5 | 9.24 | 50 | 59.5 | + | 3.88 | 0.08 | MCD | Not performed | – | – |
| IgAN-7 | IgA nephropathy | 11–15 | 11–15 | F | 102 | 4.1 | 27.1 | 18.64 | 132 | 145.4 | + | 0.5 | 0.09 | DMP | Not performed | – | – |
| Dent-1 | Dent disease type1 | 1–5 | 11–15 | F | 118.4 | 4.8 | 17.6 | 22.78 | 11764 | 77.26 | - | 0.42 | 15.23 | – | *CLCN5* | c.1562T>C | – |
| Dent-2 | Dent disease type1 | 1–5 | 11–15 | M | 130.2 | 4.7 | 19.3 | 26.33 | 15679 | 73.3 | - | 0.57 | 21.39 | – | *CLCN5* | c.800A>T | – |

| | | | | | | | | | | | | | | | | | |
|---|---|---|---|---|---|---|---|---|---|---|---|---|---|---|---|---|---|
| Dent-3 | Dent disease type1 | 6–10 | 6–10 | F | 143.3 | 4.5 | 45.3 | 89.70 | 47067 | 50.5 | - | 0.83 | 93.20 | – | *CLCN5* | c.1826T>A | – |
| Dent-4 | Dent disease type1 | 6–10 | 6–10 | F | 151.9 | 5.1 | 32.0 | 55.08 | 52418 | 58.1 | - | 0.95 | 90.22 | – | *CLCN5* | c.1826T>A | – |
| Dent-5 | Dent disease type1 | 1–5 | 6–10 | F | 70.6 | 3.9 | 419.5 | 806.42 | 104365 | 52.02 | - | 2.84 | 200.62 | – | *CLCN5* | c.1585A>C | – |
| Dent-6 | Dent disease type2 | 6–10 | 11–15 | F | 62.4 | 4.5 | 1142.9 | 931.99 | 155334 | 122.63 | - | 2.38 | 126.67 | – | *OCRL* | c.304_311del | – |
| NP-1 | Crohn's Disease | 11–15 | 11–15 | F | 124.5 | 3.1 | 1 | 1.14 | 127 | 88 | - | 0.01 | 0.14 | – | Not performed | – | – |
| NP-2 | Asymptomatic hematuria | 1–5 | 1–5 | M | 103.4 | 5 | 1 | 1.20 | 119 | 83 | - | 0.07 | 0.14 | – | Not performed | – | – |
| NP-3 | Asymptomatic hematuria | 1–5 | 16–20 | F | 96 | 4.4 | 1 | 0.54 | 63 | 186.7 | + | 0.02 | 0.03 | – | Not performed | – | – |
| NP-4 | Asymptomatic hematuria | 1–5 | 11–15 | M | 111.1 | 4.3 | 1 | 1.50 | 83 | 66.8 | + | 0.03 | 0.12 | – | Not performed | – | – |
| NP-5 | Asymptomatic hematuria | 1–5 | 1–5 | M | 140.3 | 4.3 | 1 | 1.11 | 235 | 89.7 | + | 0.11 | 0.26 | – | Not performed | – | – |
| NP-6 | Asymptomatic hematuria | 1–5 | 16–20 | F | 90.2 | 4.7 | 1 | 0.79 | 154 | 127.3 | + | 0.07 | 0.12 | – | Not performed | – | – |
| NP-7 | No abnormalities | 1–5 | 1–5 | F | 140.1 | 4.5 | 1 | 1.14 | 297 | 87.5 | - | 0.07 | 0.34 | – | Not performed | – | – |
| NP-8 | No abnormalities | 11–15 | 11–15 | F | 120 | 4.8 | 1 | 0.82 | 77 | 121.7 | - | 0.02 | 0.06 | – | Not performed | – | – |
| NP-9 | No abnormalities | 11–15 | 11–15 | F | 106.4 | 4.6 | 1 | 0.24 | 87 | 419.6 | - | 0.02 | 0.02 | – | Not performed | – | – |

PROCHOB, chronic benign proteinuria; NS, nephrotic syndrome; M, male; F, female; Cr-eGFR, creatinine-based estimated glomerular filtration rate (mL/min/1.73 $m^2$); sAlb, serum albumin (g/dL); uMB, urinary myoglobin (ng/mL); uMB/Cr, urinary myoglobin-to-creatinine ratio (ng/mg creatinine); uBMG, urinary β2-microglobulin (μg/L); uCr, urinary creatinine (mg/dL); uTP/Cr, urinary total protein-to-creatinine ratio (g/gCr); Urinary occult blood, dipstick urine occult blood (−/+); MCD, minimal change disease; FSGS, focal segmental glomerulosclerosis; DMP, diffuse mesangial proliferation. Variants are described according to HGVS nomenclature based on the following RefSeq transcripts: *CUBN* (NM_001081.4); *COL4A3* (NM_000091.5); *COL4A4* (NM_000092.5); *COL4A5* (NM_000495.5); *COQ8B*

**Supplementary Table 6. List of the genes responsible for inherited kidney diseases screened by targeted sequencing**

| | | | | |
|---|---|---|---|---|
| *SLC12A1* | *CFB* | *COL4A3* | *NUP205* | *KIRREL1* |
| *KCNJ1* | *DGKE* | *COL4A4* | *NUP85* | *SGPL1* |
| *CLCNKB* | *THBD* | *COL4A5* | *NUP133* | *LMNA* |
| *BSND* | *CFHR1* | *P3H2* | *NUP160* | *LAMA5* |
| *CLCNKA* | *ADAMTS13* | *CD151* | *CRB2* | *GAPVD1* |
| *SLC12A3* | *FN1* | *GLA* | *CUBN* | *ANKFY1* |
| *CASR* | *SLC4A1* | *UMOD* | *EMP2* | *GON7* |
| *MAGED2* | *ATP6V0A4* | *MUC1* | *FAT1* | *LAGE3* |
| *CLDN10* | *ATP6V1B1* | *SEC61A1* | *KANK1* | *OSGEP* |
| *CFTR* | *SLC4A4* | *REN* | *KANK2* | *TPRKB* |
| *CLCN5* | *CA2* | *EYA1* | *KANK4* | *TP53RK* |
| *OCRL* | *EHHADH* | *SIX2* | *PDSS2* | *WDR4* |
| *EHD1* | *SLC34A1* | *CD2AP* | *PTPRO* | *WDR35* |
| *SLC26A3* | *SLC2A2* | *NPHS1* | *XPO5* | *WDR73* |
| *KCNJ10* | *BCS1L* | *NPHS2* | *ACTN4* | *PRDM15* |
| *CLDN16* | *GATM* | *PLCE1* | *ANLN* | *TRIM8* |

| | | | | |
|---|---|---|---|---|
| *CLDN19* | *HNF4A* | *SMARCAL1* | *ARHGAP24* | *PODXL* |
| *FXYD2* | *NDUFAF6* | *LAMB2* | *INF2* | *TBC1D8B* |
| *EGF* | *CTNS* | *SCARB2* | *LMX1B* | *MAFB* |
| *TRPM6* | *NR3C2* | *COQ2* | *MYH9* | *DAAM2* |
| *TRPM7* | *SCNN1A* | *COQ6* | *PAX2* | |
| *KCNA1* | *SCNN1B* | *ITGA3* | *TRPC6* | |
| *CNNM2* | *SCNN1G* | *ITGB4* | *WT1* | |
| *HNF1B* | *KLHL3* | *GLEPP1* | *MAGI2* | |
| *PCBD1* | *CUL3* | *MYO1E* | *AVIL* | |
| *ANK3* | *WNK1* | *ARHGDIA* | *TNS2* | |
| *CFH* | *WNK4* | *ADCK4* | *DLC1* | |
| *CFI* | *AQP2* | *TTC21B* | *CDK20* | |
| *MCP (CD46)* | *AVPR2* | *NUP93* | *ITSN1* | |

**Supplementary Figure**

**Supplementary Figure 1. Urinary myoglobin decreases with improvement of proteinuria in the severe proteinuric glomerular disease group.**

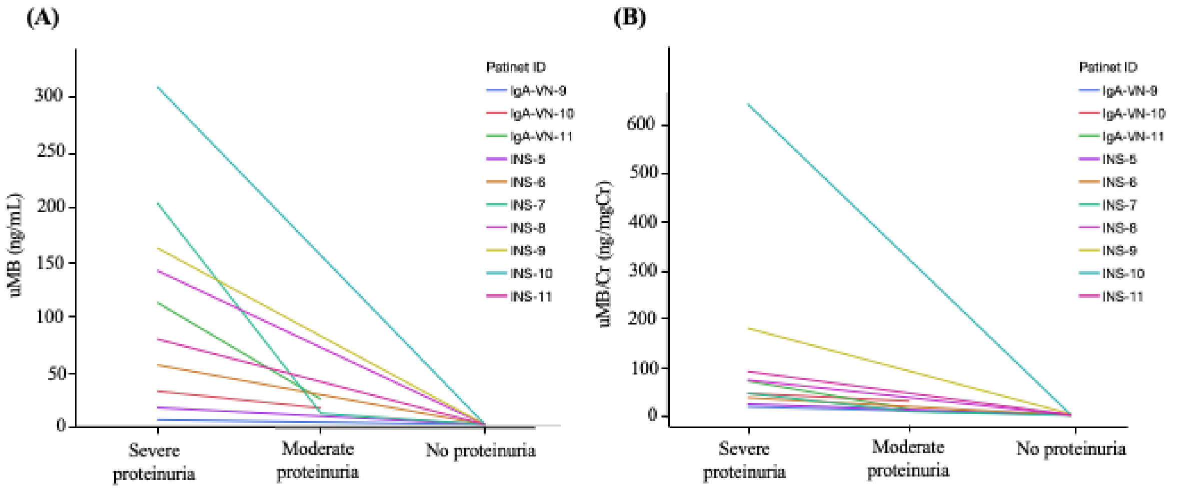


Sup-Figure 1

(A) Urinary myoglobin (uMB; ng/mL) was measured longitudinally in individual patients during treatment and is shown at up to three clinical time points: the severe proteinuria phase (urinary total protein-to-creatinine ratio (uTP/Cr; g/gCr) > 4.0), the moderate proteinuria phase (0.15 < uTP/Cr ≤ 4.0), and the no proteinuria phase (uTP/Cr < 0.15). Each line represents a single patient (patient IDs are shown in the legend).

(B) The urinary myoglobin-to-creatinine ratio (uMB/Cr; ng/mg Cr) at the same time points is shown for each patient.

**Supplementary Figure 2. Marked reduction of urinary myoglobin in samples stored in non-dedicated containers.**

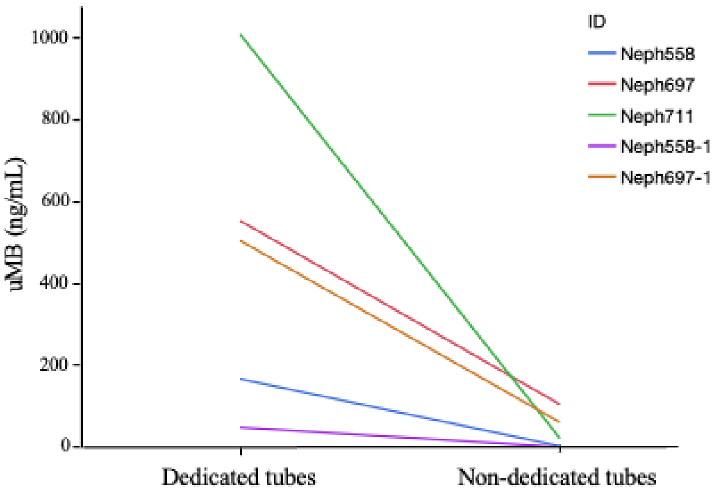


Sup-Figure 2

Urinary myoglobin (uMB; ng/mL) measured in paired frozen urine samples obtained from the same patients but stored using different containers. uMB measured from samples collected/stored in dedicated tubes was substantially higher than that measured from samples stored in non-dedicated containers. Each line represents paired measurements from an individual patient (IDs shown in the legend).
